# Airborne Transmission of Virus-Laden Aerosols inside a Music Classroom: Effects of Portable Purifiers and Aerosol Injection Rates

**DOI:** 10.1101/2020.12.19.20248374

**Authors:** Sai Ranjeet Narayanan, Suo Yang

## Abstract

The ongoing COVID-19 pandemic has shifted attention to the airborne transmission of exhaled droplet nuclei within indoor environments. The spread of aerosols through singing and musical instruments in music performances has necessitated precautionary methods such as masks and portable purifiers. This study investigates the effects of placing portable air purifiers at different locations inside a classroom, as well as the effects of different aerosol injection rates (e.g., with and without masks, different musical instruments and different injection modes). Aerosol deposition, airborne concentration and removal are analyzed in this study. It was found that using purifiers could help in achieving ventilation rates close to the prescribed values by the World Health Organization (WHO), while also achieving aerosol removal times within the Center of Disease Control and Prevention (CDC) recommended guidelines. This could help in deciding break periods between classroom sessions, which was around 25 minutes through this study. Moreover, proper placement of purifiers could offer significant advantages in reducing airborne aerosol numbers (offering orders of magnitude higher aerosol removal when compared to nearly zero removal when having no purifiers), and improper placement of the purifiers could worsen the situation. The study suggests the purifier to be placed close to the injector to yield a benefit, and away from the people to be protected. The injection rate was found to have an almost linear correlation with the average airborne aerosol suspension rate and deposition rate, which could be used to predict the trends for scenarios with other injection rates.

## I. INTRODUCTION

The transmission of the SARS-CoV-2 virus via small, exhaled airborne aerosols (*<* 5 *µ*m) has been recognized as an important pathway for the spread of COVID-19^1–6^. Smaller aerosols suspended in the air (generally termed as “droplet nuclei”) are the crystalline, virus containing, non-volatile residue left behind once the liquid in the droplet evaporates out^7–9^. These smaller aerosols could actually carry more viral load than the larger droplets, since they originate from deep within the respiratory tracts where there is more viral concentration^10,11^.

Numerical modeling (computational fluid dynamics, CFD) have been used to good effect in modeling the spread and transport of droplets via sneezing, coughing and other expiratory events^12–18^. There have been several numerical studies simulating the spread and deposition of viral droplets and aerosols in both outdoor environments^19,20^ and enclosed spaces such as hospital wards, office spaces, aircraft cabins and urinals through CFD simulations^5,18,21–31^. All these studies support the fact that ventilation, airflow streamlines, aerosol/droplet size and modes of aerosol injection are important factors affecting the transport, deposition and suspension of airborne droplets and aerosols. A recent study had investigated the effects of different aerosol source locations, particle size, glass barriers and windows using CFD simulation^32^, and it further shows how the change in airflow in the domain can significantly alter the depostion/removal patterns of the particles. Modeling of the infection spread using Monte Carlo methods^33^, and using simplified mathematical models to model the dispersion of exhaled droplets^34^ are also some important recent numerical studies of relevance to the pandemic. Studies have also attempted to perform a risk assessment in different indoor settings by studying the aerosol transport and deposition^9,24,35^, while another mathematical study has estimated the risk of airborne transmission of COVID-19 with application to face masks^36^. Facemasks and shields have been widely recommended by public health officials to reduce the spread and dispersion of viral respiratory droplets. Several recent studies have investigated the effectiveness, fluid flow behaviour and obstruction of the ejected jet through facemasks and shields.^37–42^

In classroom/healthcare settings, a high ventilation rate is required to effectively remove the airborne virus-laden aerosols from the domain. A ventilation rate of least 288 m^3^/h per person is recommended by the World Health Organization (WHO)^43^. Such a ventilation rate might not be possible to achieve through natural ventilation alone, and sometimes even in-built ventilation systems may fall short of this target. In such cases, portable purifiers might help in increasing the net ventilation rate to achieve the desired level, which needs further investigation.

Portable High Efficiency Particulate Air (HEPA) purifiers have been used for indoor purifying requirements for relatively smaller domains such as classrooms, offices and hospital wards. A few studies have studied the efficacy of air purifiers for controlling the spread of COVID-19^44–46^ and have concluded that such purifiers may serve as supplemental means for decontamination of SARS-CoV-2 aerosols. There have also been cases where portable purifiers increased the spreading of exhaled aerosols and therefore, worsened the situation^47^. Currently, there are no formal recommendations by the Center of Disease Control and Prevention (CDC) nor WHO for the usage of air purifiers. Therefore, the optimal use of air purifiers in an indoor setting remains a challenge to be studied. Our study focuses on tackling this challenge.

Spreading of the SARS-CoV-2 aerosols via wind instruments and singing cannot be ignored, as observed in a COVID-19 outbreak among a choir rehearsal group^48^. The group had followed social distancing and regulations, and yet there were 45 cases out of which two succumbed to the disease. There have been studies pertaining to the spread of coronavirus through aerosols ejected from wind instruments, although most of them focused on the airflow from the instruments^49–51^. A recent study examined the aerosol generation from different wind instruments and quantified the risk for each instrument^52^. Singing can also be a dangerous source of virus-laden aerosols, having an injection rate typically greater than normal breathing and speaking^53–55^. Both singing and wind instrument playing can take place in a music classroom, prompting the need for careful consideration of the safety regulations and protocols inside these classrooms.

This study examines the effects of portable air purifiers inside a music classroom, which are placed at different locations to determine the most strategic placement. In addition, this study determines whether adding a purifier does help in improving the ventilation and the airborne aerosol removal times from the domain. Moreover, this study also examines different injection modes (such as using musical instruments, singing and normal breathing) with different injection rates for each mode (e.g., with and without masks, different instruments). The airborne aerosol concentration at the elevations of interest, the deposition of aerosols onto the surfaces inside the domain, and the amount of aerosols filtered by the purifiers are some key findings which will be reported in this study.

## II. NUMERICAL MODELING

### A. Computational fluid dynamics (CFD) simulation framework

The simulations are conducted based on the CONVERGE CFD platform version 2.4^56^. The Eulerian-Lagrangian framework is used for the gas-aerosol simulation. CONVERGE uses a nearest node approach to exchange mass, momentum, energy terms of a parcel (Lagrangian particle) with the fluid-phase (Eulerian field) values of the computational node that it is closest to. A Taylor series expansion is used to calculate the gas velocity (Eulerian field) at the point of the parcel (Lagrangian particle). The use of the Taylor series expansion significantly reduces grid effects on the spray. A collocated finite volume approach is used to numerically solve the conservation equations. Flow quantities are calculated and stored at cell centers according to the summed fluxes through the cell faces and an internal source term, if any. The gas phase flow is governed by the conservation equations of mass and momentum. The incompressible form of the equations are given below (since the Mach number is very low and the gas density is close to constant):

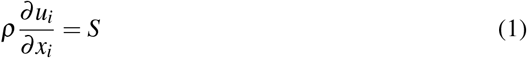

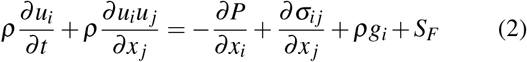

where *σ*_*ij*_ is the viscous stress tensor given by:

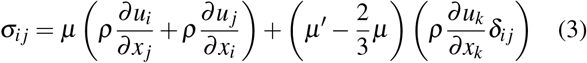

where *t* is time, *g*_*i*_ and *u*_*i*_ are the gravitational acceleration and velocity component in the *i*-th direction, respectively, *ρ* is the density of the gas, P is the pressure, *µ* is the viscosity, *µ* is the dilatational viscosity (set to zero) and *δ*_*ij*_ is the Kronecker delta function. *S* and *S*_*F*_ are the source terms incurred by the Lagrangian particles, which are calculated by:

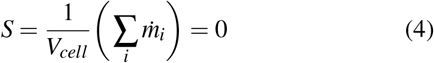

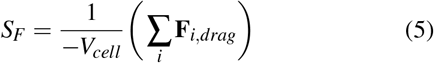

where the summation over *i* means the summation over all the Lagrangian particles within a cell, 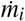 is the rate of change in mass of a particular Lagrangian particle (this term is zero since break-up is neglected, and evaporation is assumed to have already reduced the particles to the minimum size: see more details below) and **F**_*i,drag*_ is the drag force on the Lagrangian particles. Evaporation is turned off for the duration of the simulation, since it was assumed that all the droplets ejected from the musical instruments (which are already *<* 5 *µ*m in size^52^) quickly evaporate to the minimum size (chosen to be 1.5 *µ*m in this study), as justified in Shao *et al*.^9^. Aerosols around 1.5 microns are essentially the crystalline, non-volatile components leftover when the liquid in the droplet evaporates out. The assumption that the small droplets quickly evaporate into droplet nuclei within an order of few seconds is justified in recent and past studies^7,9,14,15,20,57,58^. Figure 1 reproduced from the study of Morawksa^7^ shows how the droplets starting at 1 *µ*m and 10 *µ*m evaporate to the residual size within 10 milliseconds and 1 second, respectively (even at up to 80% relative humidity). These small 1 *µ*m particles also take around 30,000 seconds to completely fall to the ground via natural gravitational settling^7^. Hence, the deposition of these particles are largely affected by the airflow in the domain, and the initial droplet size distribution does not matter as long as it is within the O(10 *µ*m) range, because the evaporation will drive them to the crystalline size within a second.

The dispersed Lagrangian particles are modeled as spherical, 1.5 *µ*m particles with a density equal to that of air at 300 K (*ρ* = 1.161 kg/s). This assumption is validated by the fact that droplet nuclei leftover from fast evaporation possess very little inertia and hence follow the airflow^7^. This assumption is also supported by a recent study^59^, where the authors found that gravity and inertia play little role on particles *<* 10 *µ*m. In addition, the ejected aerosols are pretty dilute (around 500 - 2000 particles per liter of ejected airflow) based on experimental observations^9,52^ and hence, the interactions between Lagrangian particles are also ignored. The mass rate of change 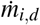 and the total force (acting on the particle) **F**_*i,d*_ govern the dynamics of each Lagrangian particle by:

**FIG. 1:**
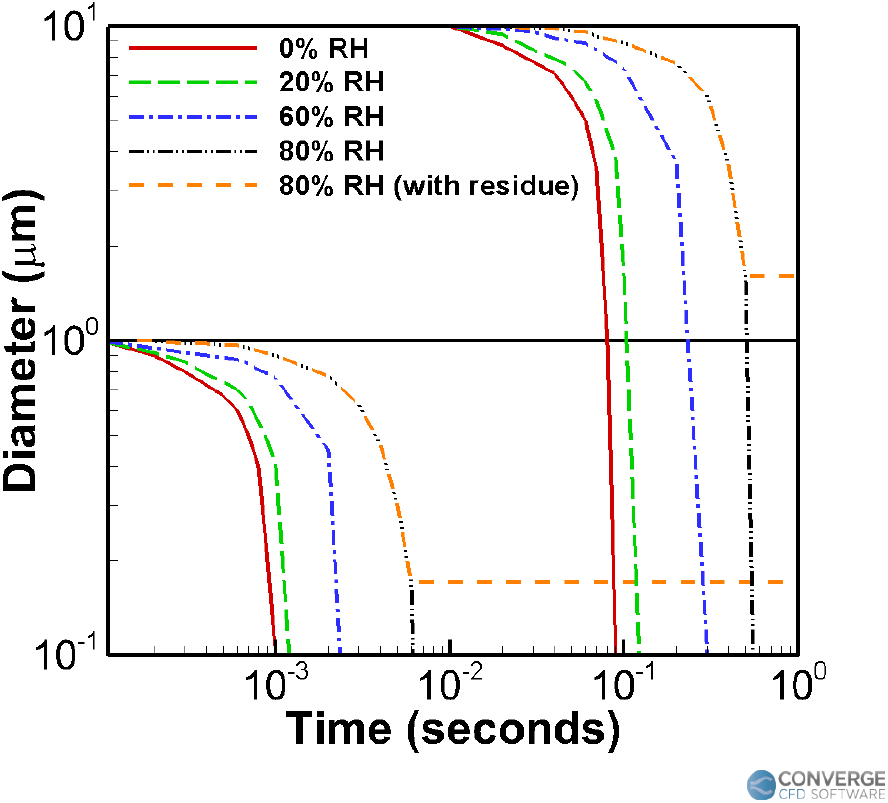
Changes to water droplet diameter as a result of evaporation, taken for two different initial droplet sizes (1 and 10 *µ*m) and for different conditions of relative humidity (RH), as shown in the study of Morawksa^7^.

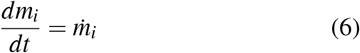

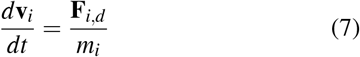

and

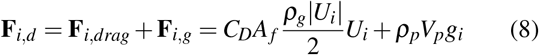

where *C*_*D*_ is the drag coefficient, *A*_*f*_ = *πr*^2^ is the particle’s frontal area (and *r* is the radius of the particle), *ρ*_*g*_ is the gas density, *ρ*_*p*_ is the particle density, *V*_*p*_ is the particle volume, and *g*_*i*_ is the gravitational acceleration in the *i*-th direction. *U*_*i*_ is the particle-gas relative velocity in the *i*-th direction given by:

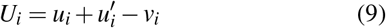

where *u*_*i*_ and 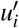 are the local mean and turbulent fluctuating gas velocities in the *i*-th direction, respectively. Equation 7 can be expanded as follows:

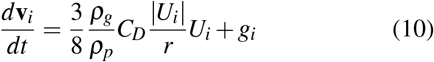

Only the Stokes drag is considered for the drag force^60^, since the small aerosols are close to spherical shape and fixed in size (no distortion or break-up). It is vital to incorporate the effects of turbulent fluctuations and particle dispersion in these flows^13,20,34,61,62^. The Reynolds Averaged Navier-Stokes (RANS) turbulent simulations are conducted with the *k* − *ε* model^63^ for the Eulerian gas-phase flow, along with the O’Rourke turbulent dispersion model^64^ for the Lagrangian particles.

### B. Geometry and computational mesh

The confined space of the classroom contains both the student as well as the teacher, or only the student depending on the case. The geometry details were obtained from the University of Minnesota (UMN) School of Music. This classroom is frequently used for one on one tutoring sessions or solo practise sessions for the students, and is hence very vital to the school. The orientation of the domain is shown in Fig. 2a, the labeled objects (piano, student, teacher, inlets and outlets) are shown in Fig. 2b, and the locations are shown in Fig. 3. The humans are 1.6 m tall (with the injectors located around 1.5 m high), with the aerosol injection being either from an wind instrument (a trombone or a trumpet), or directly from the nose. The instrument has an outflow diameter of 10 cm and a length of 50 cm. In the singing case (Fig. 3a), the singer’s mouth is 4 cm in diameter. For the piano case (Fig. 3b), the injection cavity (which is the nose during normal breathing) is from a 1.25 cm diameter orifice.

**FIG. 2:**
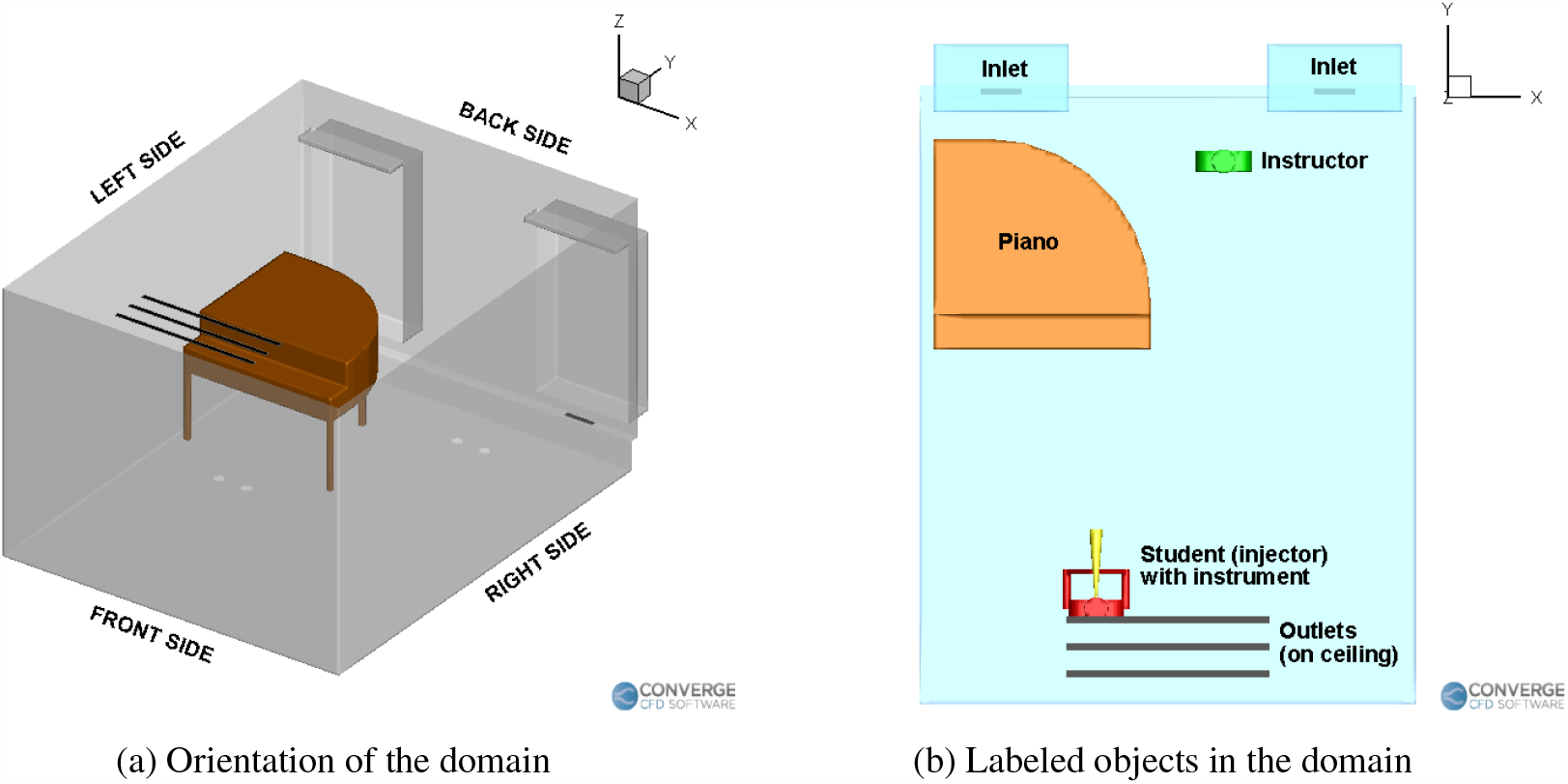
Orientation and objects in the domain.

**FIG. 3:**
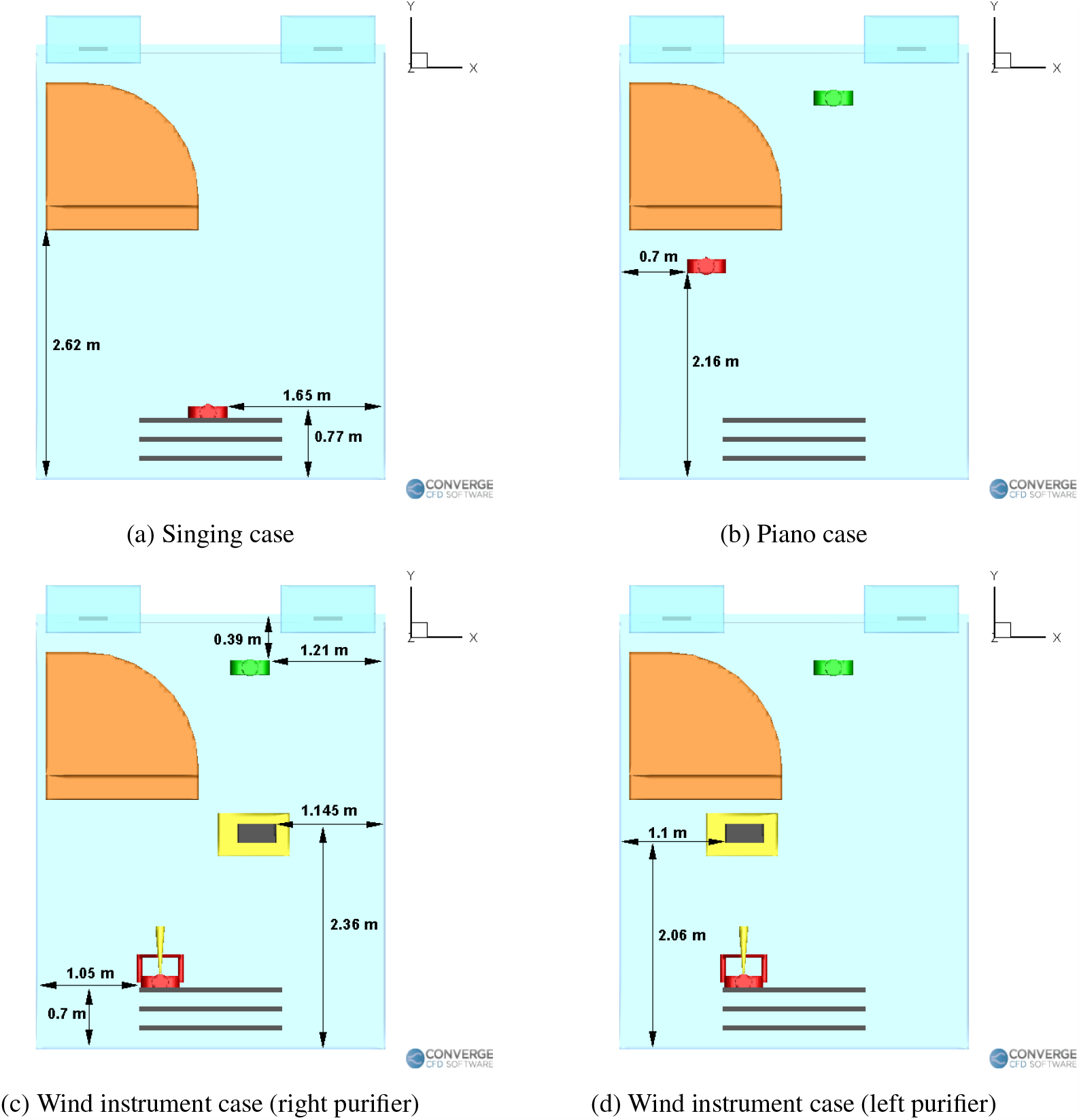
Locations of the student, teacher and purifier in the different scenarios.

Table I lists all corresponding dimensions of the domain parameters. CONVERGE uses a numerically stable cartesian mesh and employs a unique cut-cell approach that perfectly represents the underlying geometry as provided by the user. Mesh refinement has been applied at certain boundaries (such as the inlets and outlets), as well as the particle ejection region in front of the aerosol emitter. The total volume of the domain is 45.8 m^3^. A base grid size of 0.05 m was used at the more open areas of the domain, while smaller grid sizes of 0.0125 m and 0.025 m were used in the refined regions. The total number of cells were 400,000. A minimum time step of 0.005 s was used (a variable time step algorithm keeps the time step within 0.005 – 0.01 s). Around 384 CPU hours were used for a simulation of 11 minutes.

**TABLE I:** The dimensions of important objects in the domain.

| Object/<br>surface | Dimensions [m]<br>X - Y - Z |
| --- | --- |
| Room | 3.657 - 4.472 - 2.743 |
| Inflow vents | 0.3 - 0.05 |
| Outflow vents | 1.5 - 0.05 |
| Piano | 1.55 - 1.55 - 1.2 |
| Purifier | 0.4064 - 0.2032 - 0.638 |
| Table | 0.75 - 0.45 - 1.0 |
| Human | 0.4 - 0.15 - 1.6 |

### C. Boundary conditions

The no slip boundary condition is applied at all solid surfaces (except the vents). Aerosols stick to the boundaries upon contact. At the ventilation air inlets, a constant mass flow rate of 0.05663 kg/s (which corresponds to an Air Change per Hour, ACH, of 3.63, or a Cubic Feet per Minute, CFM, of 100) is applied, with a zero normal gradient for pressure. These values were obtained from the technicians in the UMN School of Music. At the air outlets, a zero normal gradient condition was applied for velocity, while a constant outlet pressure of 1 atm was specified. The temperature was specified to have a uniform value of 300 K throughout the domain. The purifiers have a constant suction rate of 0.1046 kg/s (190 CFM or an ACH of 6.76), based on the CFM specifications of a standard Fellows AeraMax 209 purifier^65^.

## III. RESULTS AND DISCUSSION

The simulation results of the different case settings are presented and discussed in this section. Two major effects are examined in this study, namely the effect of an air purifier (specifically, its improvement in ventilation and the effect of its location), and the effect of different aerosol injection rates. There are three types of musical sessions taking place: 1) a student singing alone in a room; 2) a student playing a wind instrument inside the room with the teacher present; and 3) a student playing the piano with a teacher present in the room.

For the singing cases, the simulation time is 2,160 s, with 600 s of injection and 1,500 s of idle time. The initial minute of pure airflow is present for the singing cases as well. For the wind instrument and piano cases, the simulation time is 660 seconds (with an initial minute of pure airflow).

### A. Effect of purifiers

This section examines the impact of placing purifiers in the domain. As will be seen, introducing purifiers affects the ventilation rates and airflow streamlines in the domain, leading to changes in the number of aerosols remaining in the air, deposited onto surfaces and removed from the domain. Moreover, the effect of placing the same purifier at different locations in the domain is also examined in this section.

#### 1. Comparison with CDC/WHO guidelines: Improvement in aerosol removal times due to increase in ventilation rates

Two key points are studied: Firstly, the effects of introducing a purifier into the domain are analyzed by comparing the profiles of airborne, deposited and removed aerosols between the benchmark case without a purifier (for the singing case) and the case with a purifier. Subsequently, the ventilation rates and aerosols removal times (by introducing a purifier) are compared with established WHO & CDC guidelines.

The student (located exactly at the center underneath the return vents, see Fig. 3a) is singing inside an empty room for a duration of 10 minutes. The initial 1 minute of simulation was conducted with pure airflow from the vents, and with no aerosol injection. This was to let the airflow field develop into a “statistically stationary” state, after which the aerosols are subsequently injected. After the 10 minutes of singing, there is then a break for 25 minutes (during which time the student is not present in the room) and the airborne aerosols are allowed to settle and deposit, such that the room can be safer before the next person’s arrival. It is of interest to examine the number of remaining airborne aerosols in the room. The rate of aerosol injection is 700 aerosols/s^55^, with an airflow rate of L/s^53,66^. It is of interest to note that singing has a slightly larger particle injection rate (700 aerosols/s) when compared to normal speaking (570 aerosols/s) as found in Alsved *et al*.^55^. For the purifier case, a purifier is placed on the ground in front of the piano. The purifier is a Fellows AeraMax 290 model with a CFM of 190 (around 0.1046 kg/s or an ACH of around 6.76). One assumption we make is that the purifier removes all the viral aerosols when passed through the HEPA filter. This assumption is justified, since HEPA filters are required to have at least 99.97% (or higher) removal efficiencies for particles larger than 0.2 *µ*m^44^.

Figure 4 shows the airflow streamlines for the singing case without a purifier. The airflow streamlines flow from the right side of the room to the left side (Fig. 4b), and it can be seen that the piano obstructs the flow on the left. This causes recirculation zones near it (Fig. 4b and Fig 4a), causing deposition to occur near the regions around the piano. Figure 5 shows the streamlines inside the room for the singing with a purifier case. We can see that the recirculation zones forming on both the vertical plane (going over and below the piano, Fig. 5a) and the horizontal plane (going from the right side to the left side of the room, Fig. 5b). These streamlines are slightly different from the case where there was no purifier, which is expected, since the airflow rate of the purifier is higher than the existing building HVAC ventilation airflow rate. The purifier therefore drives the airflow streamlines in the domain, especially in the region near the injector.

**FIG. 4:**
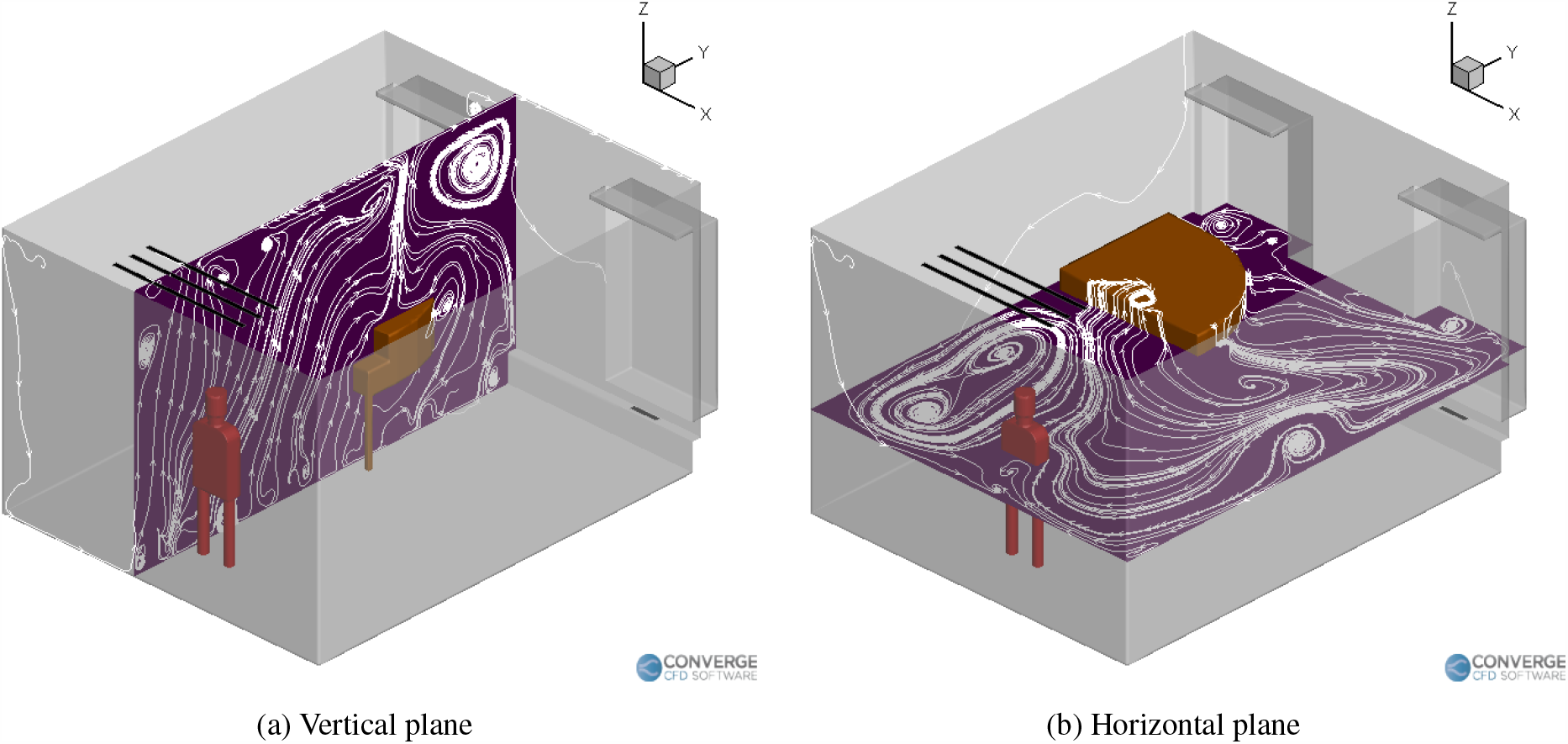
Airflow streamlines inside the room with a singer but without a purifier.

**FIG. 5:**
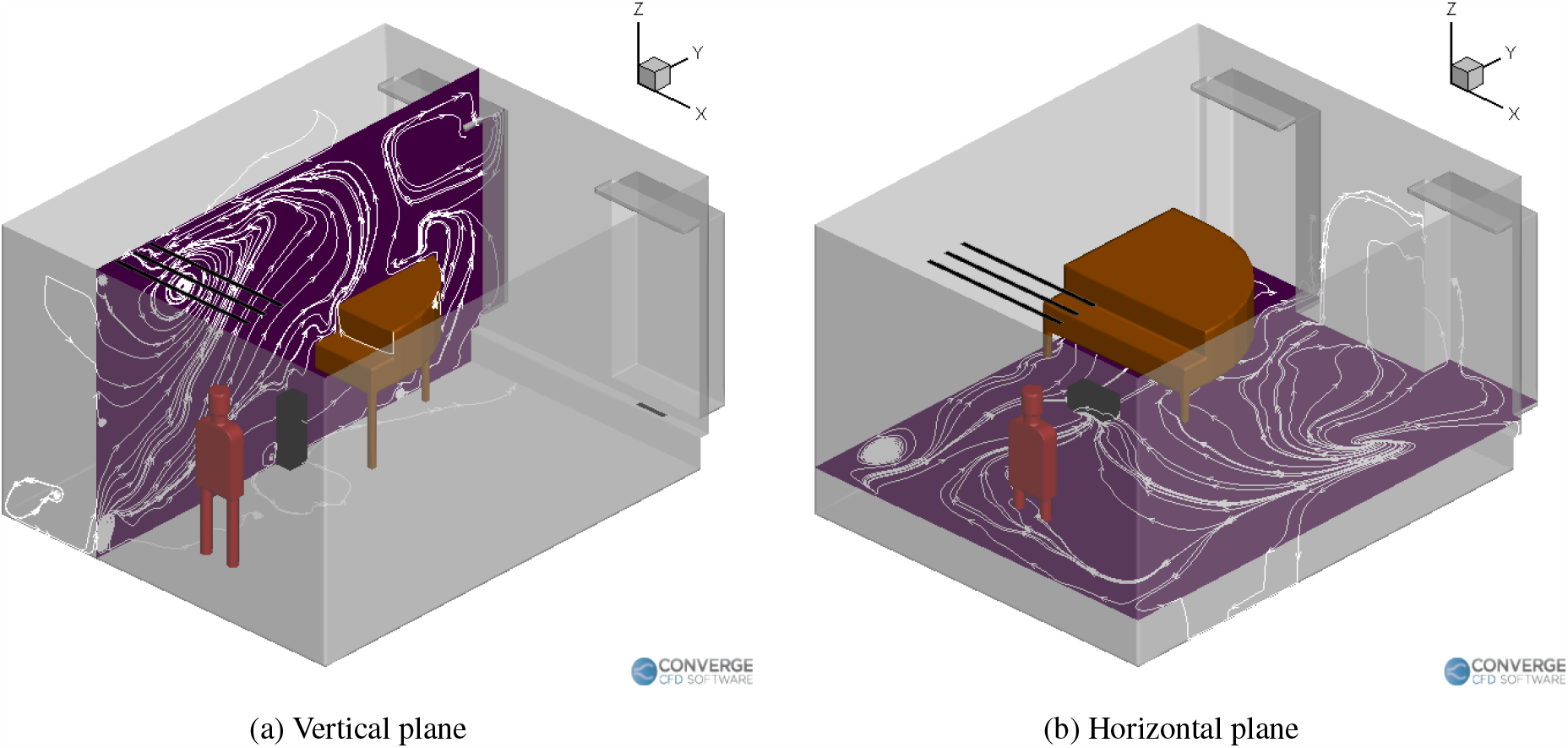
Airflow streamlines inside the room with a singer and a purifier.

Figure 6 compares the deposition of the aerosols in the domain between the singing with no purifier case (Fig. 6a) and singing with a purifier case (Fig. 6b) at the end of 36 minutes. Much of the deposition occurs on and near the piano, especially underneath the piano. Significant deposition occurs on the vent strip (containing the air inlets), as well as on the window sills. Additionally, there seems to be some deposition on the student themselves, which is an important point to consider. The clothes of the students need to be well washed to prevent further risk of spreading to others. It is observed that for the purifier case, there is slightly less deposition near the left side walls and window. The purifier increases deposition near the ground in front of it, causing less deposition on the ground near the back side of the piano. Overall, the deposition trend seems to be similar between the two cases. This result dictates which surfaces of the room need to be cleaned thoroughly, especially the regions on, and around the piano.

**FIG. 6:**
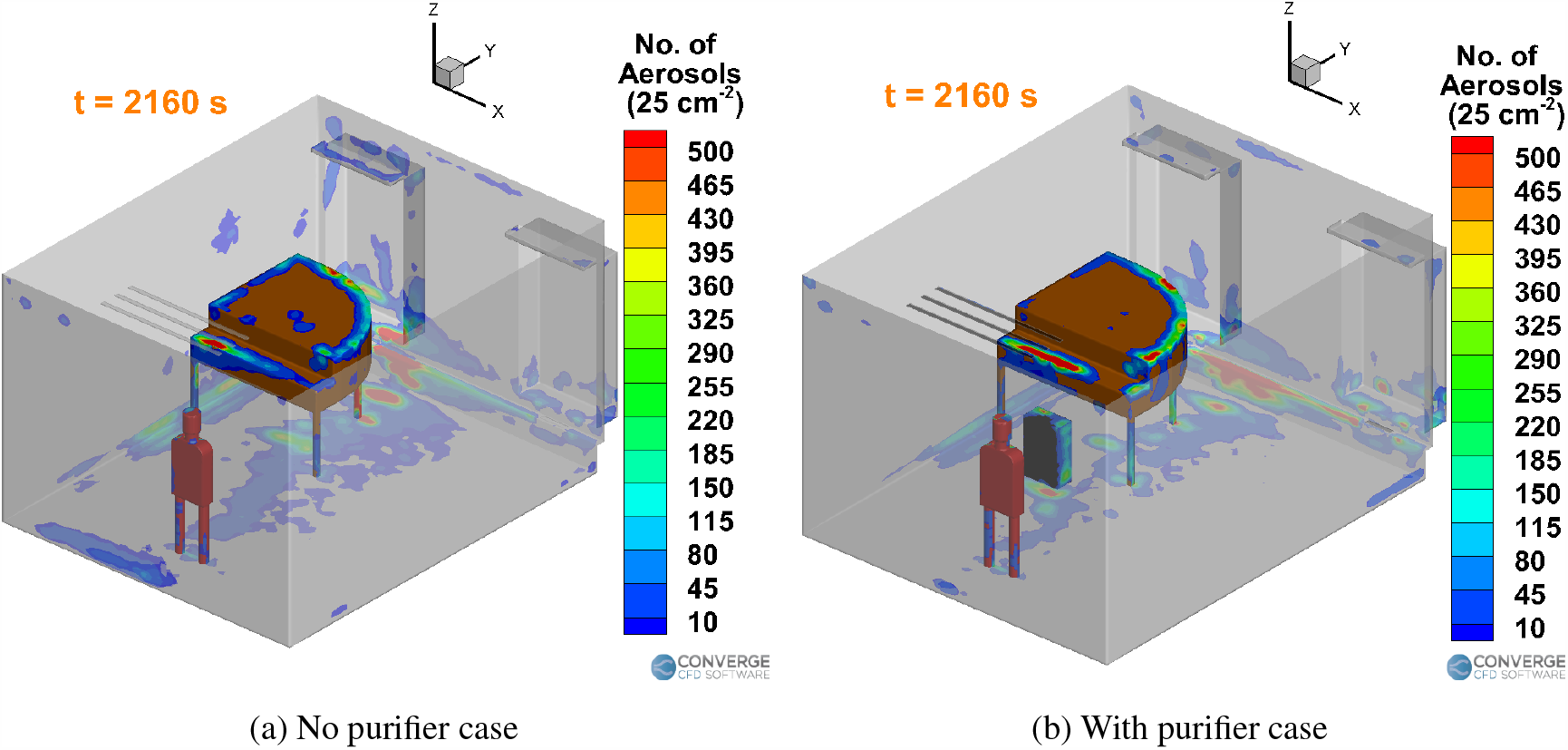
Aerosol deposition per unit cell area (25 cm^2^) inside the room with a singer.

In general, we can first conclude that the airflow stream-lines inside a domain significantly affect the transport of the aerosols. Moreover, the presence of any large objects which can obstruct the flow of the inlets (in this case, the piano), will alter the streamlines drastically. Obstructing the inlet airflow would lead to recirculation zones near these objects, which can cause significant deposition of aerosols near those objects. Regions which are fairly free from such large objects (in this case, the right side of the room) do not experience significant deposition. Hence, deposition here is being affected by the change in the airflow streamlines due to the geometry, as well as the multiple airflow inlets (including the instruments and purifiers) and outlets.

Figure 7 summarizes the effect of having a purifier in the singing case. For both the singing cases, the number of airborne aerosols fluctuates about a mean value, at around the 500 to 600 second mark. The singer stops singing at the 11 minute mark, and leaves the room for the break. The total number of aerosols injected into the domain in 10 minutes of injection time is 420,000. Figure 7a compares the time varying profiles of airborne aerosols in the domain between the with and without purifier cases. The number of airborne aerosols rises until the 500 to 600 second mark, to a peak value of around 111,943 (26.6% of the total number of aerosols injected in the domain, which is 420,000) for the no purifier case and 114,888 (27.3%) for the purifier case. Subsequently, the airborne aerosol number drops rapidly and reaches a near-steady value of 8,699 (2%) for the no purifier case at the end of 36 minutes. Further reduction in airborne aerosol numbers is extremely slow, around ∼ 1 aerosol per 10 s. This is because the aerosols have been stably trapped in recirculating streamlines, thus leading to almost no further drops in aerosol numbers. For the purifier case, the corresponding airborne aerosol number value at the end of 36 minutes is around 4,333 (1%), but unlike the no purifier case, this value is still decreasing at a rate of 2 to 5 aerosols/s. This is because the purifier’s presence continues to drive some removal to occur via the streamlines going through it. Figure 7b compares the time varying surface deposition number profiles between the two cases. The number of deposited aerosols steadily increases and levels off at a value of around 411,108 (97.8%) for the no purifier case and 386,611 (92%) for the purifier case, at the end of 36 minutes. The deposition is thus lessened slightly for the purifier case, although the difference is minor (around 6% variation from the no purifier case). The time varying removal of aerosols for the two cases is compared in Fig. 7c. The number of aerosols removed in the no purifier case is around 168 (0.04%), while the corresponding number is around 29,006 (7%) for the purifier case. Of these, around 16,000 (3.8%) aerosols were removed by the purifier itself, and around 13,000 (3.2%) were driven out through the ceiling outlets (Fig. 7d). Thus, the total number of aerosols removed from the domain for the case with a purifier (around 29,006) is more than two orders of magnitude higher than the case without a purifier (around 168).

**FIG. 7:**
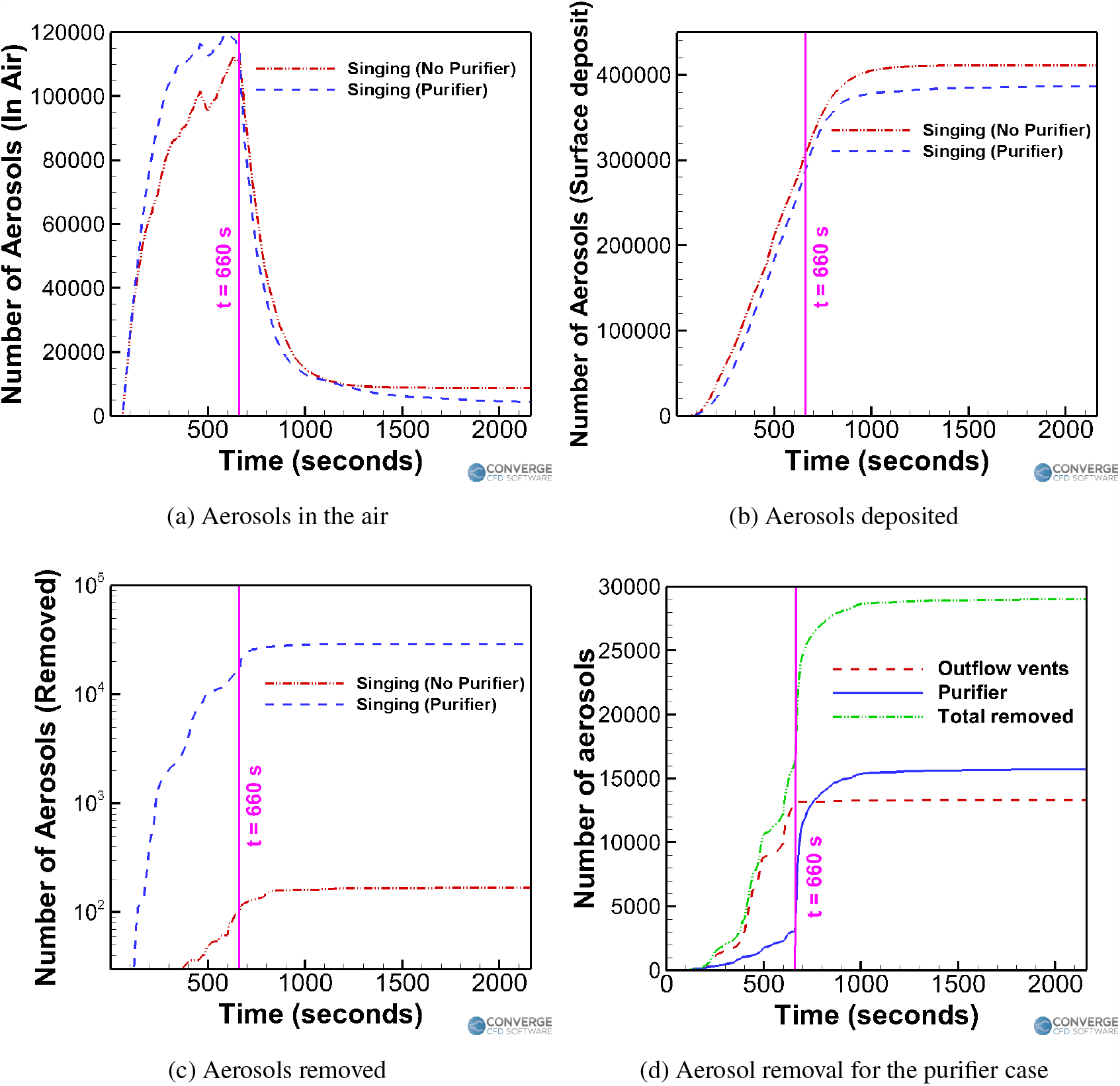
Trend comparison for the singing cases.

When not using any purifier, the existing building HVAC airflow rate of 0.05663 kg/s corresponds to a ventilation rate of around 166 m^3^/h, which is significantly less than the ventilation rate of least 288 m^3^/h per person is recommended by WHO^43^. However, adding a purifier of 0.1046 kg/s (which by itself corresponds to around 322 m^3^/h) increases the overall ventilation to around 488 m^3^/h, which is far more than the required rate prescribed by WHO.

Improvements in ventilation rates by adding a purifier also helps in reducing removal times. The CDC guideline-suggested levels of removal times, are shown in Table II, reproduced from the study of Chinn *et al*.^67^. The table is valid for a room without any continuous injection, which corresponds to our singing cases (where the student stops singing and leaves the room at the 11 minute mark).

**TABLE II:**
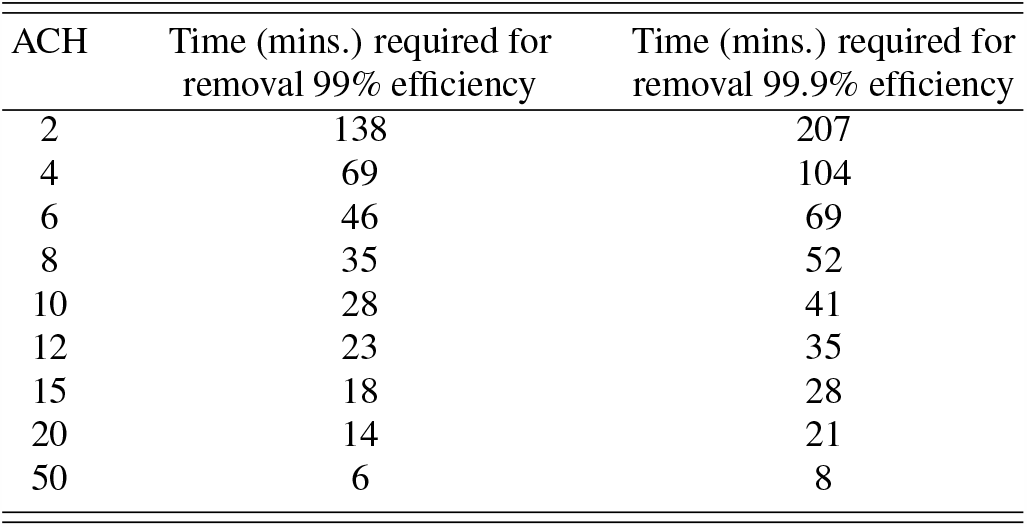
CDC guidelines for the Air changes/hour (ACH) and time required for airborne-contaminant removal by efficiency^67^.

For the case without a purifier, if we expect a slow removal rate of 1 aerosol/10 s (assuming a best case), it would take around 24 hours (i.e, an entire day) to completely remove the remaining aerosols. According to the CDC guidelines for a room with an ACH of 3.63, it should roughly take around 81 minutes for 99% removal and around 123 minutes for 99.9% removal. Clearly, these numbers are very far off from the expected removal time of *>*24 hours. This suggests that the natural aerosol removal capacity of the classroom is not sufficient enough to effectively remove all (or most) of the airborne aerosols.

For the case with a purifier, if we assume a steady airborne aerosol removal rate of 1 aerosol/s (which is less than what was observed at the 36 minute mark), it would take around 53 minutes more (in addition to the 25 minutes duration of no injection) to reach the 99% removal stage, where only 1,149 aerosols are remaining airborne (1% of the 114,888 airborne aerosols left when the singer stops singing). It would also take 70 minutes more to reach the 99.9% removal stage, where only 115 airborne aerosols remain. Considering that 25 minutes have already elapsed since the singer stopped singing, it would take roughly 78 minutes and 95 minutes for achieving the 99% and 99.9% removal efficiency respectively, which are faster than the CDC guideline values of 81 minutes and 123 minutes, respectively.

Table III summarizes the aerosol removal times of the cases with and without a purifier, with the CDC guidelines. In conclusion, adding a purifier helps in improving the ventilation rate to a value above the recommended rate prescribed by WHO, while also helping in achieving aerosol removal times as prescribed by the CDC guidelines. Moreover, this helps in deciding an effective break period in between sessions of singing/instrument playing. The break period of 25 minutes used in the singing case with a purifier reduces the airborne aerosols to almost 4,333 (from 114,888), which is a reduction of almost 97%. This could serve as an effective break period even for lengthened periods of any classroom session, since it is observed that the number of airborne aerosols fluctuates about a mean value after an initial transient period of around 8 to 9 minutes (for the singing case).

**TABLE III:** Comparison of the removal times (with and without a purifier) to the CDC guideline values.

| ACH | Time for 99% removal efficiency |  |  | Time for 99.9 % removal efficiency |  |  |
| --- | --- | --- | --- | --- | --- | --- |
|  | CDC | Purifier | No Purifier | CDC | Purifier | No Purifier |
| 3.63 | 81 min | 78 min | > 1 day | 123 min | 95 min | > 1 day |

#### 2. Effect of purifier location: impact on airflow streamlines, aerosol deposition and removal

In the previous section, the benefits of adding a purifier were examined. Here, we observe the how the trends (aerosol deposition, airborne aerosol numbers and aerosol removal) change when the purifier is placed at different locations. In this scenario, the student is playing a wind instrument (trombone), with the teacher present at the opposite end of the room. The aerosol injection rate is 30 aerosols/s^52^, with an airflow rate of 600 mL/s from the instrument^68^. As in the previous singing case, the initial one minute of simulation was conducted without any aerosol injection, and with just pure airflow from the vents and purifiers. Aerosols are then injected for 10 minutes, after which the simulation stops. Purifiers are switched on right from the start of the simulation, and run throughout the entirety of the session.

Firstly, the immediate benefit of introducing a purifier (on the ground, similar to the singing case) can be observed visually from Fig. 8, which compares the aerosol clouds at a few instances of time between a case without a purifier, and a case with a purifier. In both cases, the aerosol cloud shifts to the left side of the room due to the airflow streamlines in the domain (as will be seen later in Fig. 10 and Fig. 14), which leads to more deposition near the piano. The piano also obstructs the flow from the left side inlet and causes changes in the airflow streamlines in all cases. We can see a visible reduction in the airborne aerosol cloud and the deposited aerosols for the case with a purifier. But the question arises if this location for the purifier is an optimal one.

**FIG. 8:**
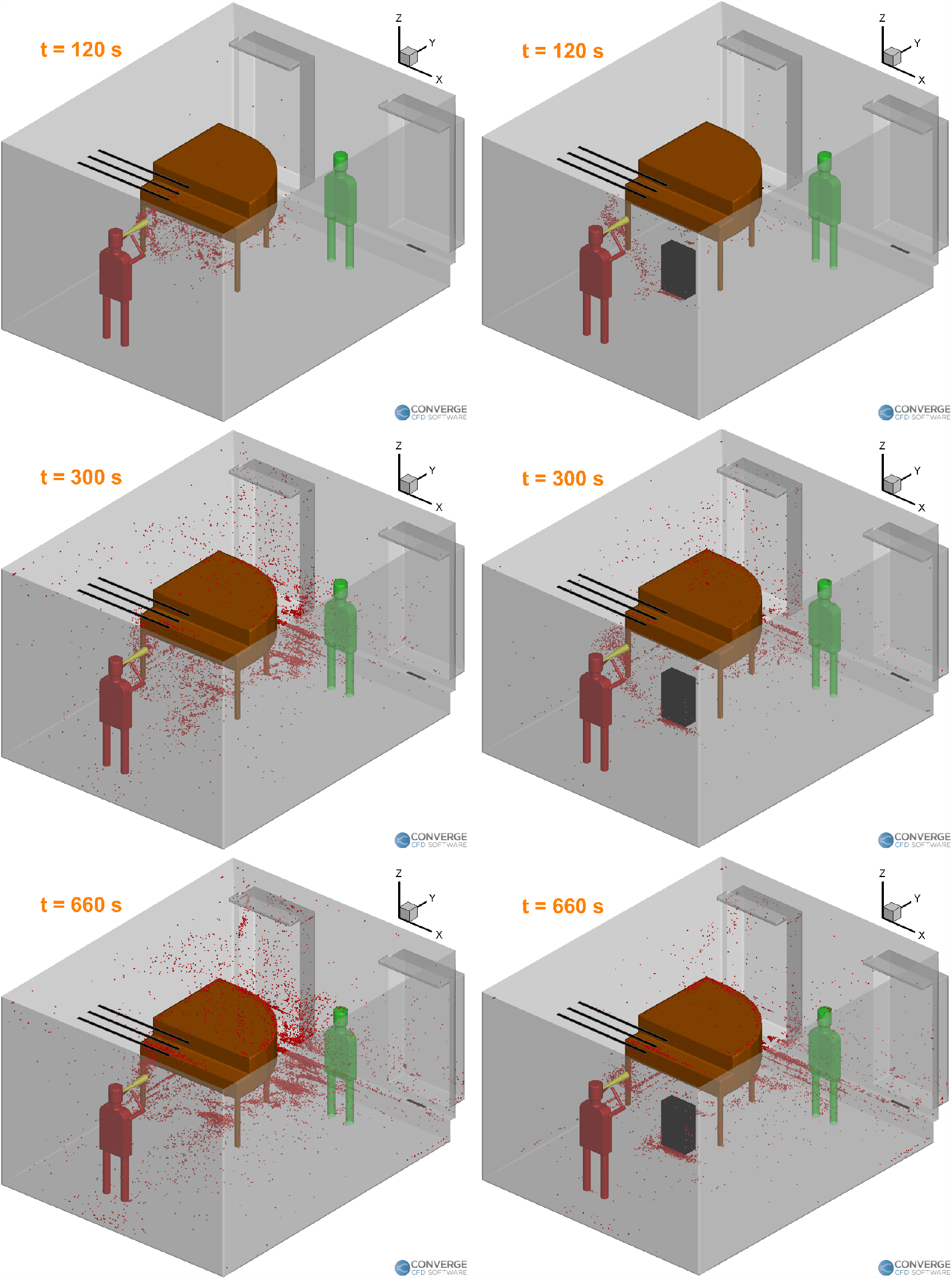
Aerosol cloud profiles at three instances of time inside the room: left side without a purifier; right side with a purifier on the ground.

Therefore, three different purifier arrangements are analyzed - purifier placed on a table on the left, purifier placed on a table on the right, and a purifier placed on the ground on the left. The elevations (achieved by using a table) are 1 m high. In order to draw meaningful conclusions, these three arrangements are compared to a case without any purifier.

Figure 9 compares the deposition profiles of the aerosols onto the surfaces of the domain in the four cases (three purifier arrangements, and the case without a purifier). The deposition pattern of the case without a purifier (Fig. 9a) and the case with an elevated purifier on the left (Fig. 9b) are very similar to each other. There seems to be a slight reduction in deposition for the purifier on the ground case (Fig. 9c). However, the purifier on the right side case shows a drastically different deposition profile (Fig. 9d), where most of the deposition is limited to the edge of the walls near the piano.

**FIG. 9:**
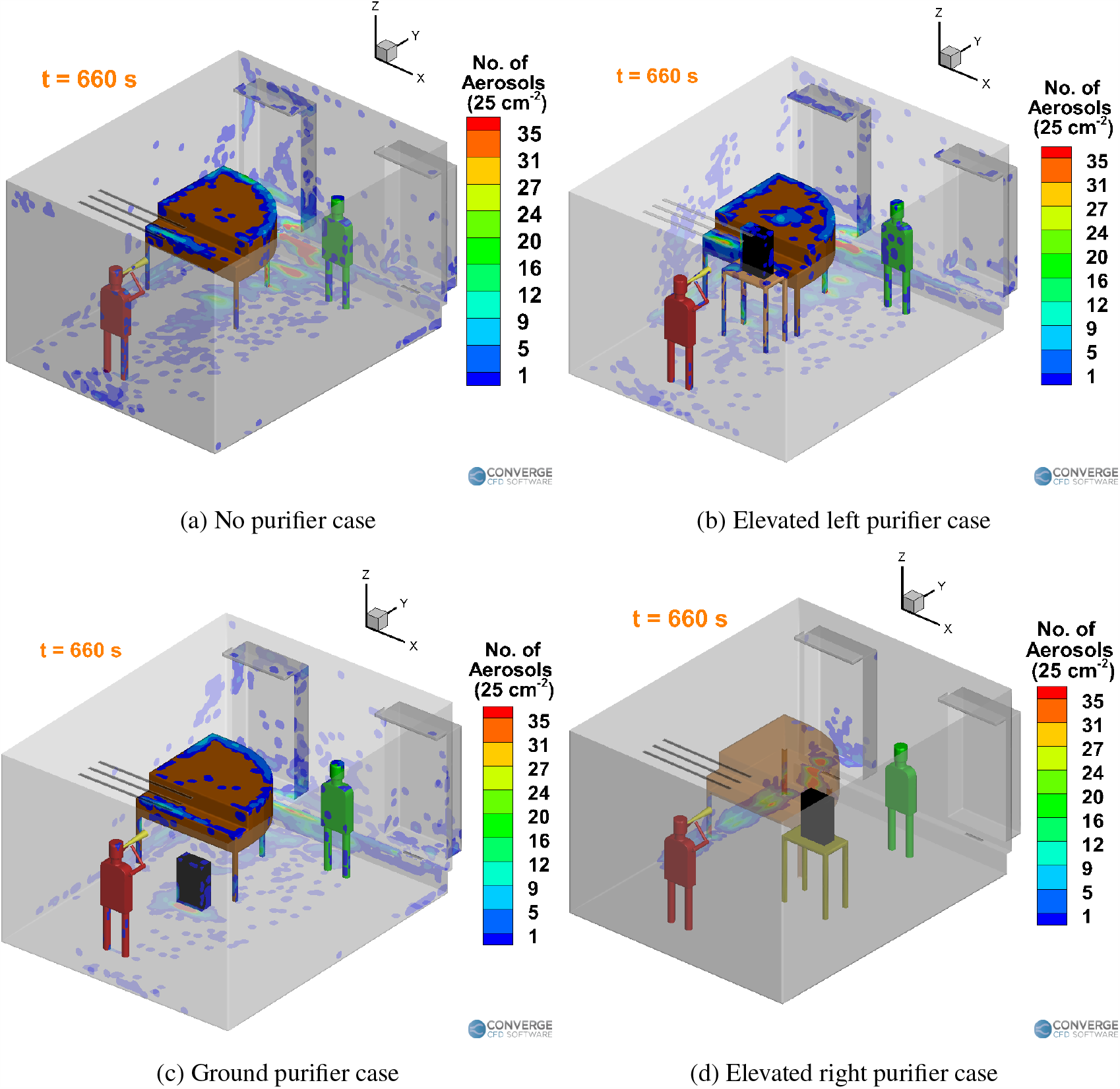
Deposition trends for the wind instrument (trombone) case.

The deposition patterns can be explained by the airflow streamlines in the room. The presence and location of the purifier largely affects the airflow streamlines, which in turn affect the deposition and airborne concentration of the aerosols. The airflow streamlines for the case without a purifier is shown in Fig. 10. The average velocity magnitude in the room is around 0.1 - 0.2 m/s (see Fig. 11a) at the injector elevation (1.4 m), except near the room inlets (0.3 m) where the velocity is around 1.6 m/s (see Fig. 11b) and near the piano top, along with the area near the windows directly above the inlets (around 0.4 m/s, see Fig. 11a).

**FIG. 10:**
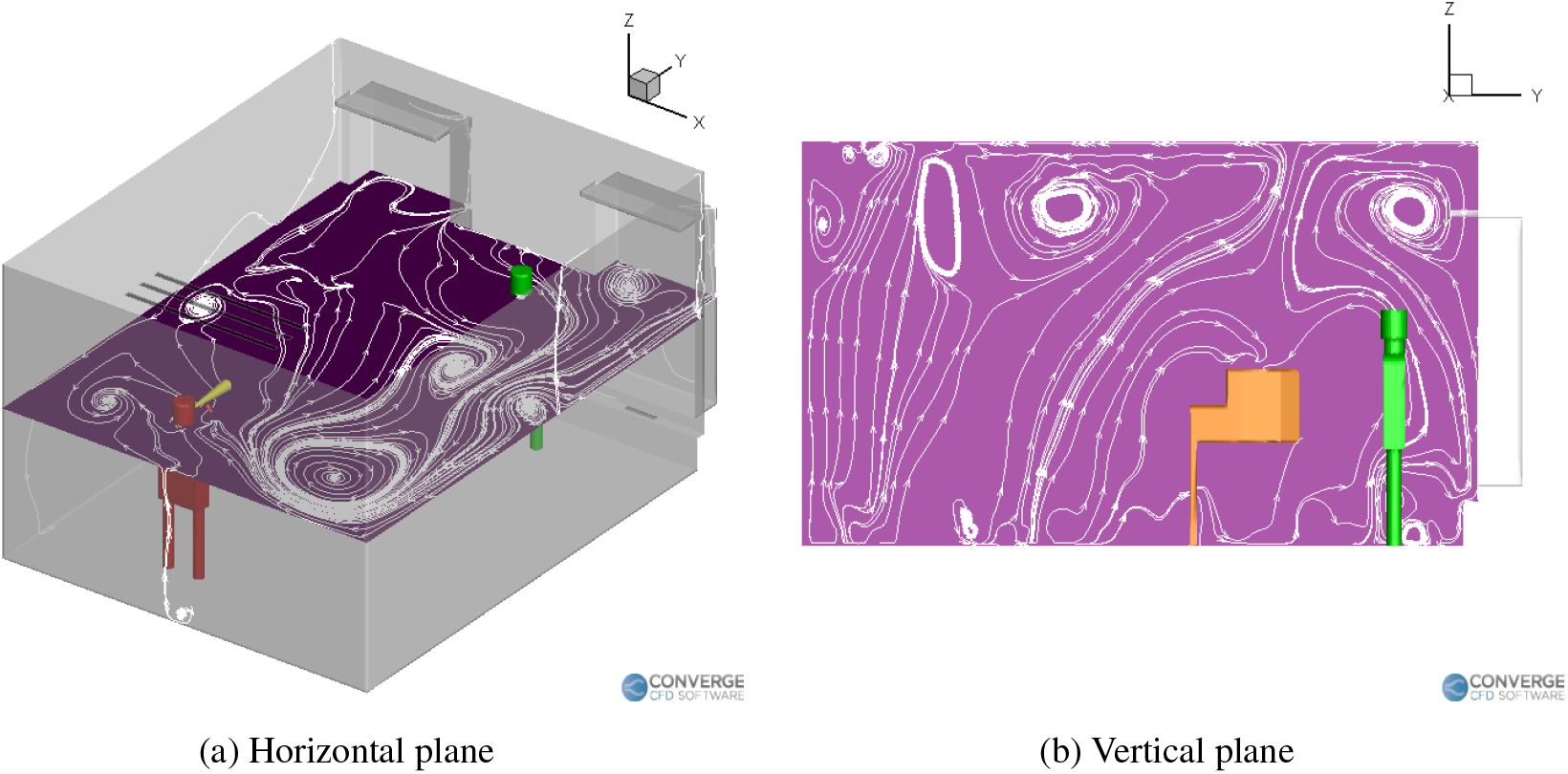
Airflow streamlines inside the room (no purifier case).

**FIG. 11:**
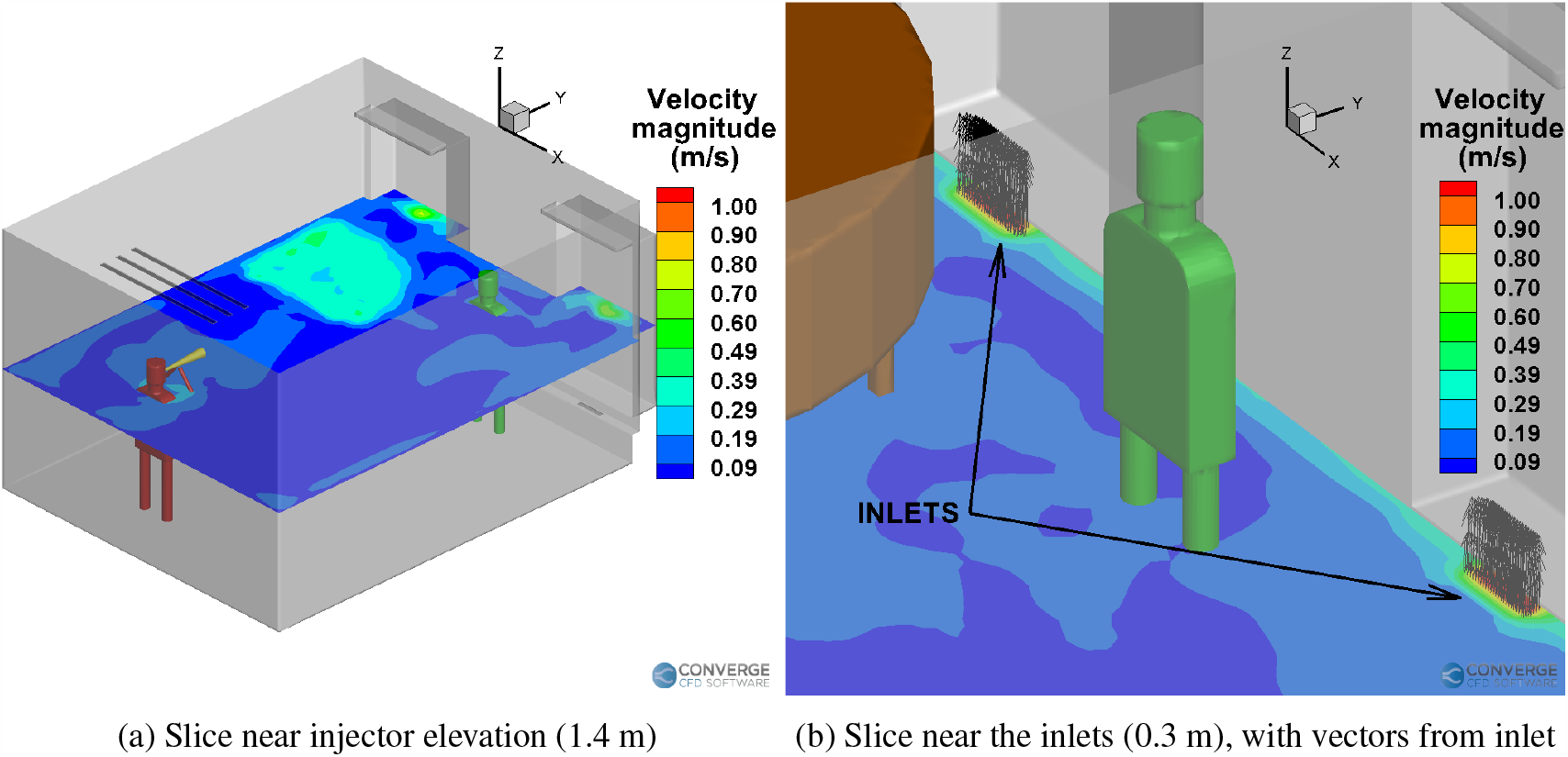
Time averaged velocity magnitude contours for the no purifier case.

For the elevated left side purifier case (Fig. 12), the purifier’s suction side directs the airflow streamlines towards the center of the room (Fig. 12a), while the exhaust of the purifier quickly blows away the aerosols towards the rear side of the room and towards the teacher (see Fig. 12a and Fig. 12b). This “boost” zone behind the purifier serves to accelerate the deposition of aerosols on top of the piano, as seen in the deposition profile (Fig. 9b). Figure 13 shows the airflow streamlines in the room due to the presence of the purifier on the right side of the room, at an elevation. Two sets of slices are taken to show the difference in streamlines near and away from the injector location, as the purifier is far away from the injector in this case. We can immediately see that the presence of the purifier on the right side drastically changes the streamline profiles. The horizontal recirculation zones no longer extend all the way from the right side to the left side, as seen in the previous cases (Fig. 10 and Fig. 12). Instead, the streamlines divert towards the center-right portion of the room, where the purifier is located (Fig. 13c). The aerosols at this elevation (near the injector) subsequently tend to flow to lower heights due to the weaker airflow velocity on the left side. At the lower elevations where the purifier’s influence is weaker, the streamlines do extend all the way to the left side of the room (Fig. 13d). Due to this, the aerosols tend to deposited onto the extreme left side of the wall once they settle to the low elevation. The horizontal streamlines for the purifier on the ground case (Fig. 14a) also extend from the right side to the left. Moreover, the exhaust of the purifier directs the airflow underneath the piano (Fig. 14b), which then flow upwards behind the piano to create a strong recirulation zone there, which causes aerosols to deposit near the center of the piano (Fig. 9c). The purifier on the left table case has a similar recirulation zone on top of the piano (Fig. 12b), but it is smaller than the recirculation zone seen in Fig. 14b. We can see some of the streamlines flowing right onto the center of the piano, which causes some deposition near the center of the piano (Fig. 9b).

**FIG. 12:**
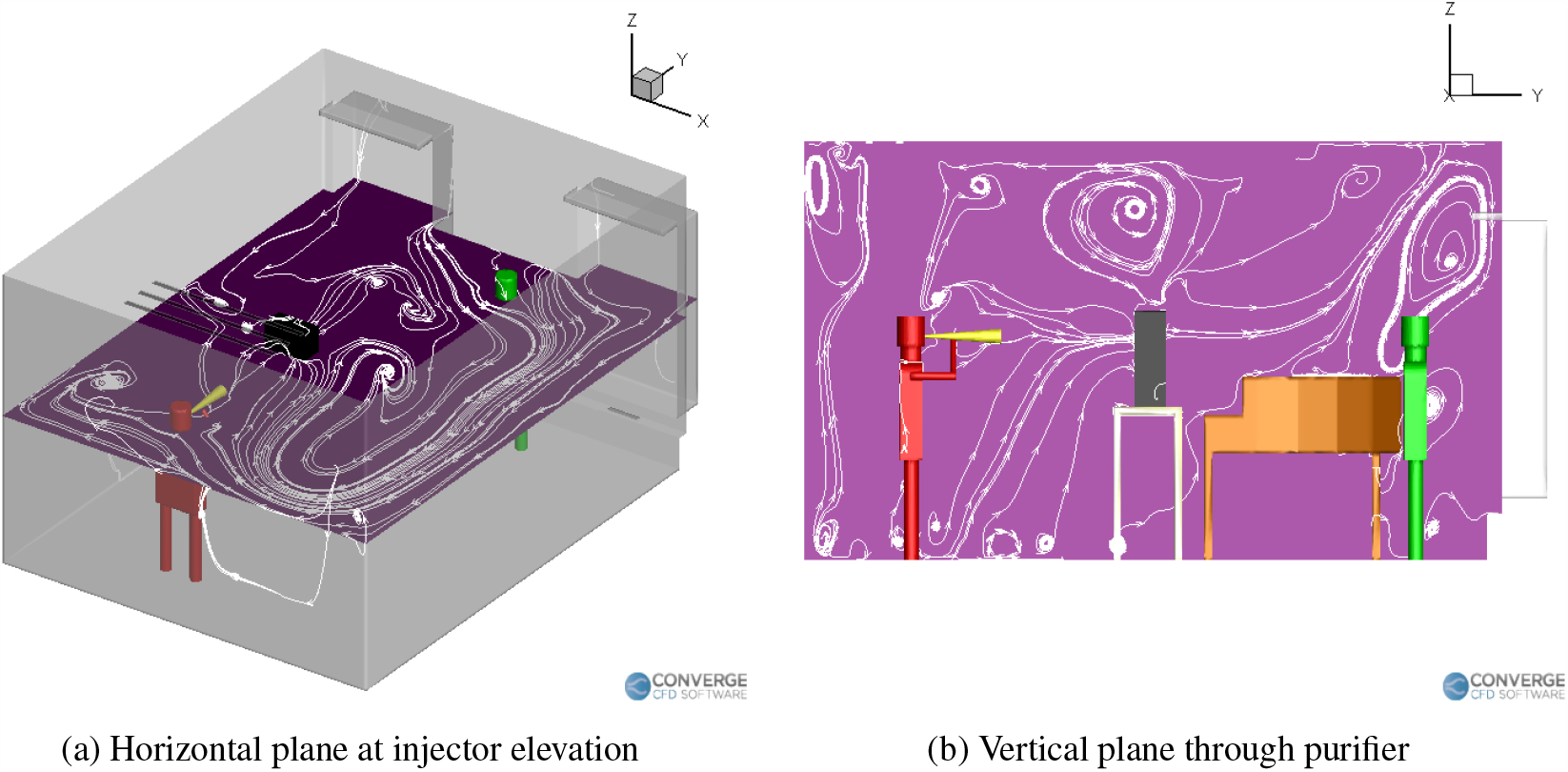
Airflow streamlines inside the room with a purifier on the left table.

**FIG. 13:**
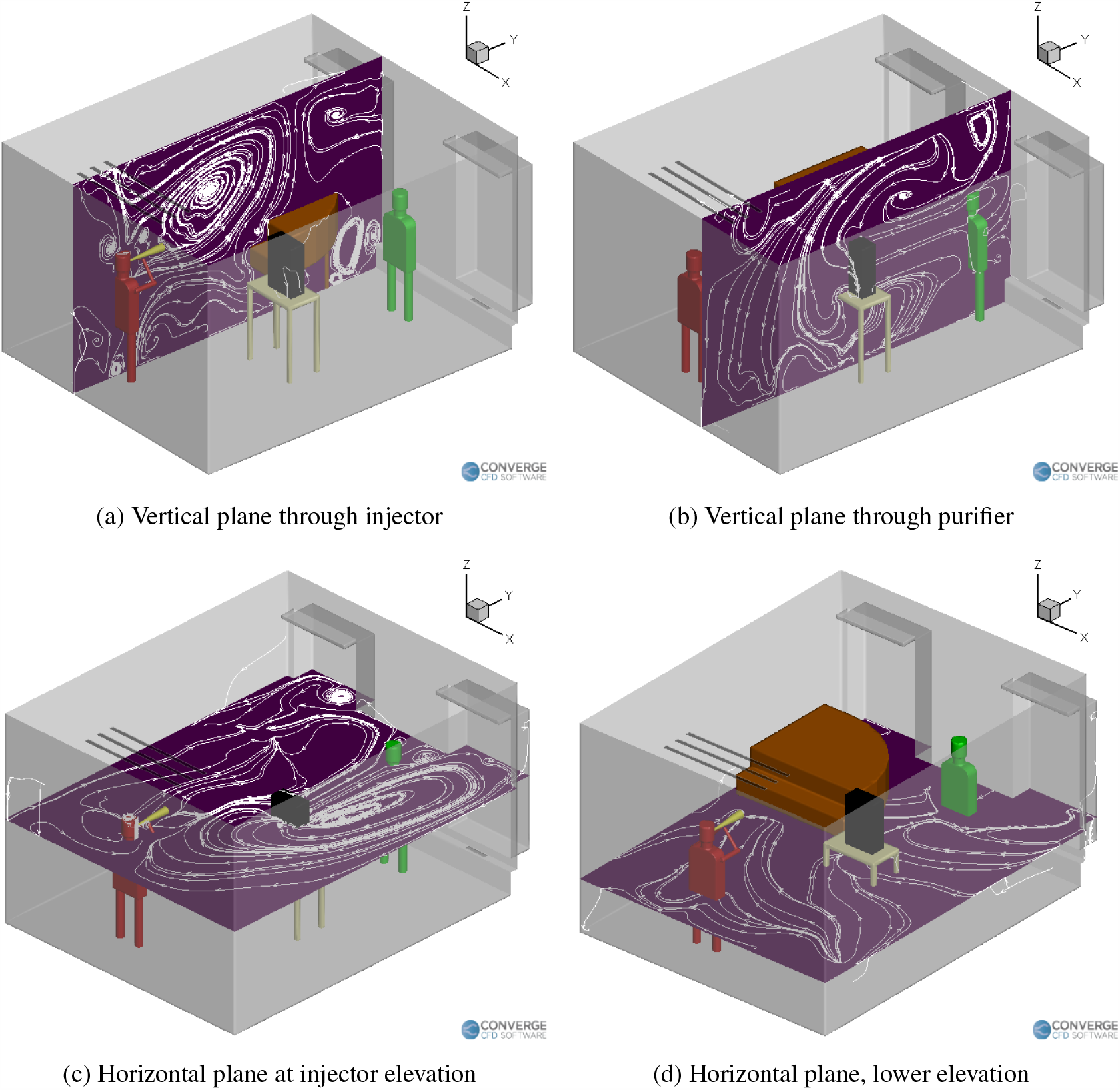
Airflow streamlines inside the room with a purifier on the right table.

**FIG. 14:**
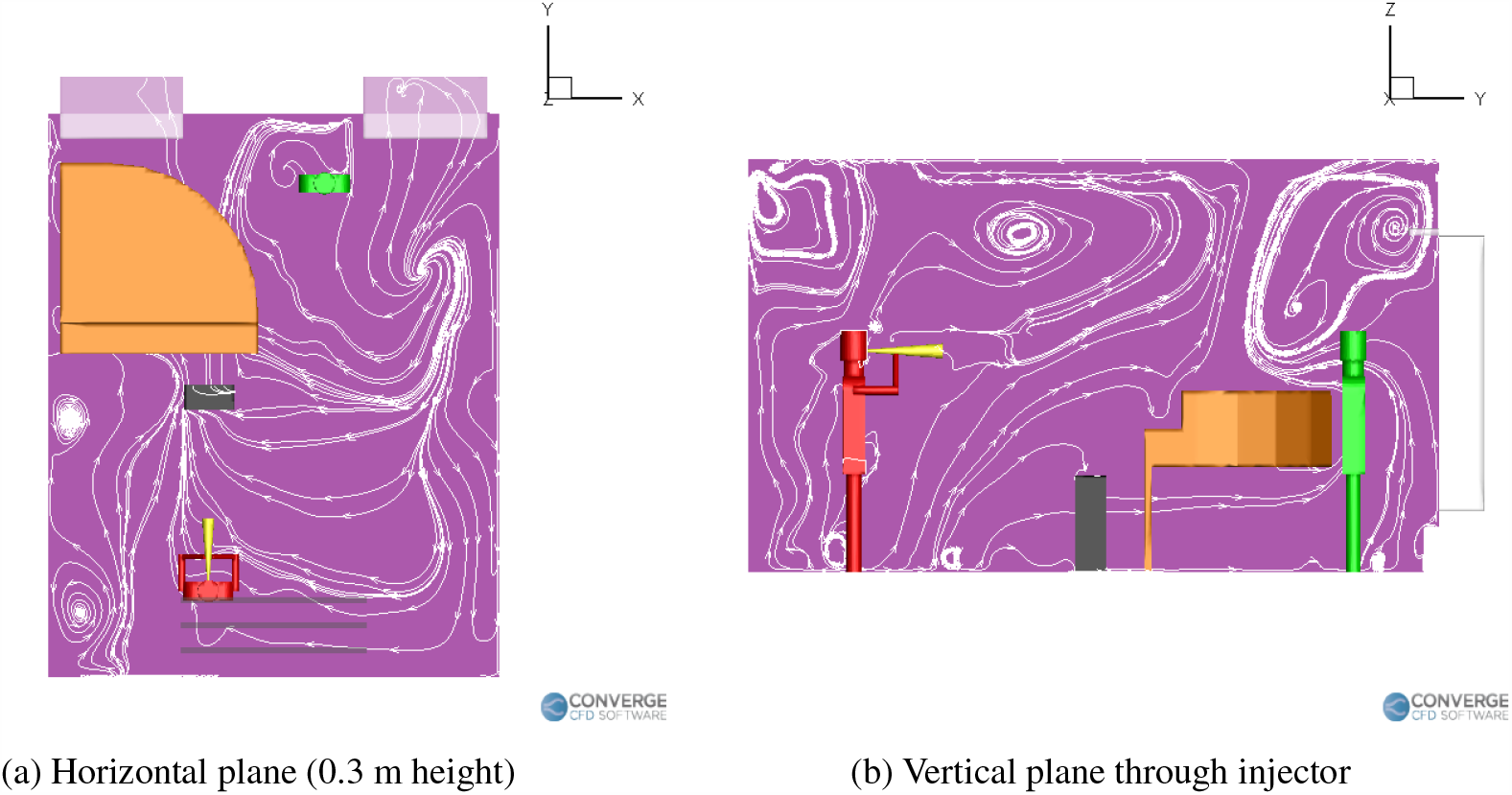
Airflow streamlines inside the room with a purifier on the ground.

In general, we can comment that purifiers are significantly going to affect the airflow streamlines in the room, which in turn will affect both deposition, the airborne concentration profiles, as well as the removal of the aerosols through the vents/purifiers. It is recommended to place the purifier in a location which does not disrupt the natural airflow streamlines in the room in a negative way (the exhaust of the purifier should not cause more mixing/spreading of the aerosols). An example of proper purifier placement can be found in the purifier on the ground case, where the airflow streamlines from the exhaust of the purifier follow a path underneath the piano (Fig. 14b), which is similar to the case without a purifier (Fig. 10). Hence, there is not significant spreading of aerosols in this case. On the other hand, in the purifier on the left side case, the purifier’s exhaust causes airflow streamlines above the piano (Fig. 12b) which causes more mixing and hence, slightly more deposition onto the teacher (Fig. 9b).

Figure 15 compares the temporal profiles of the number of aerosols among the different purifier cases with the no purifier case. It is observed that for all these cases, the number of aerosols remaining in the air more or less oscillates about a mean value after an early transient period (observed to be around 5 to 6 minutes). Any simulation time greater than the initial transient period will produce results which will vary in values, but not in trend behavior. Further injection of aerosols after this point primarily causes the deposited number to grow, with little increase in the number of airborne aerosols. A total of 18,000 aerosols injected into the domain during the simulation duration. From Fig. 15a, a final airborne aerosol number of 3,951 (22.2%) is observed for the benchmark (no purifier) case. Compared to this, the number of airborne aerosols has reduced to 3,219 (17.9%) for the elevated left purifier case, and even lower to 1,534 (8.5%) for the ground purifier case. Interestingly, the number of airborne aerosols has actually increased to 9,093 (50.5%) for the elevated right purifier case, making it more dangerous than the case without a purifier (in terms of airborne aerosol numbers). As for the number of aerosols deposited onto surfaces (Fig. 15b), the benchmark case exhibits a final deposition number of 14,049 (77.8%). The deposition numbers for the purifier cases are 11,249 (62.5%) for the elevated left purifier case, 8,465 (47%) for the ground purifier case and 8,907 (49.4%) for the elevated right purifier case. This shows that the ground purifier case has consistently reduced both the airborne aerosol number as well as the deposited aerosol number. In order to determine the number of aerosols removed from the domain in each case, we take a look at Fig. 15c. The benchmark case exhibits close to zero removal of aerosols, suggesting that the current building HVAC flow rate is insufficient to remove any aerosols from the classroom. The elevated right side purifier case also exhibits close to zero aerosol removal which, when combined with its higher-than-benchmark airborne aerosol number, makes it the worst performing purifier case. The elevated left side purifier case exhibits a removal of 3,530 (19.6%), while the ground purifier case exhibits a removal of 8,001 (44.4%). Thus, the grounded location of the purifier has served to be the best one so far. It lies naturally in the path of the ejected aerosols, and is also more isolated from the teacher due to it being hidden away underneath the piano. As a result, it offers the best reduction in airborne and deposited aerosol numbers compared to the no purifier case, while also exhibiting the highest aerosol removal number among all the cases.

**FIG. 15:**
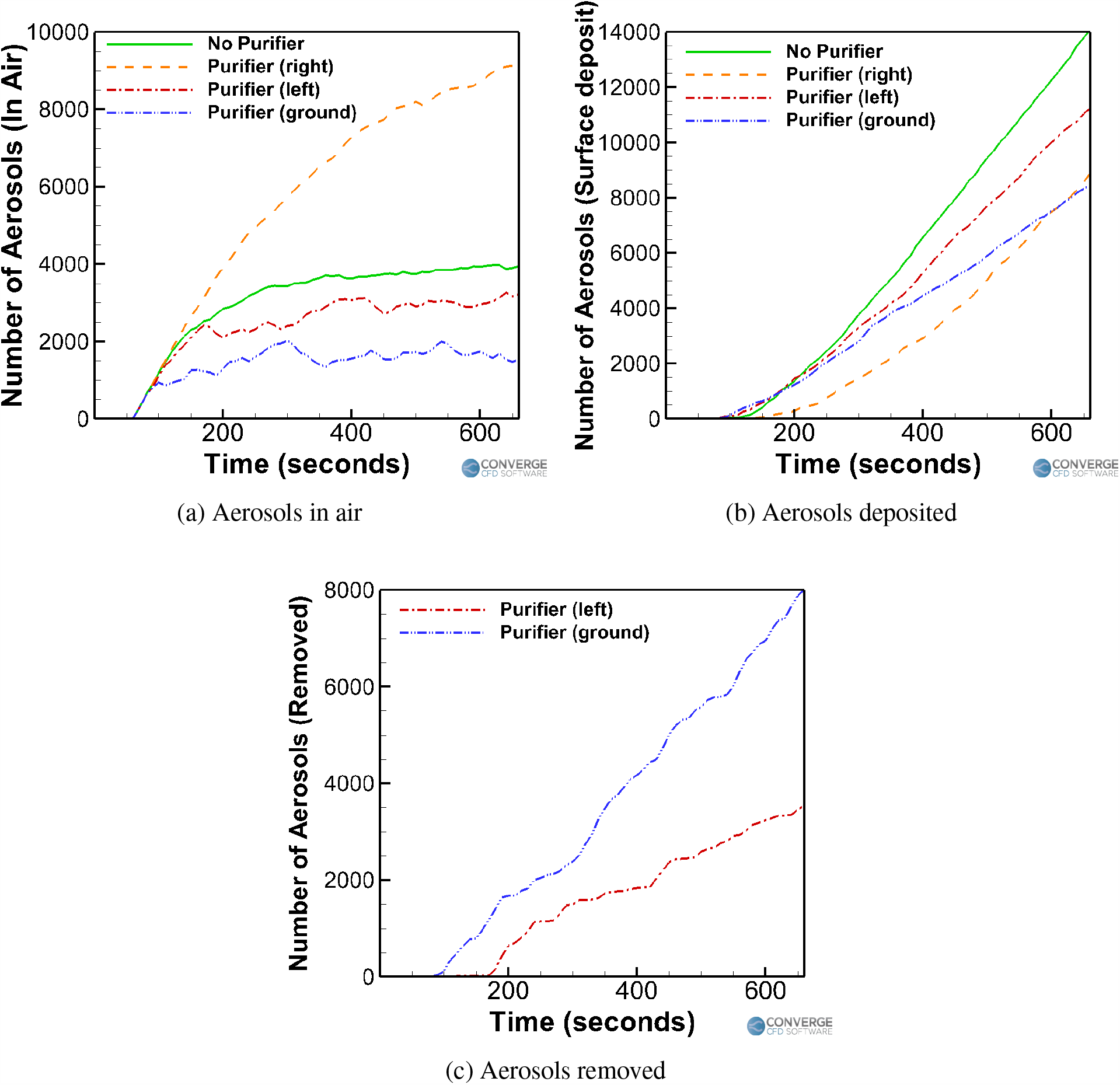
Trend comparison for the wind instrument (trombone) cases.

Figure 16 compares the time averaged airborne aerosols concentration at elevations of interest, inside the classroom between the no purifier benchmark case (Fig. 16a and Fig. 16b) and the remaining cases with purifiers. In Fig. 16a, the Z-plane slice is located at a height of 0.5 m from the ground, while the Y-plane is located at a distance of 4.1 m from the front of the room. A fairly widespread region of aerosol presence is observed underneath the piano, as well as behind the piano. This is due to the airflow streamlines carrying the aerosols through these regions, as shown in Fig. 10. In Fig. 16b, the Z-plane slice is located at a height of 1.4 m, while the Y-plane slice is located at a distance of 4.0 m from the front of the room (this almost coincides with the plane of the teacher’s nose and mouth). There is a region of aerosol presence above the piano, owing to the streamlines and recirculation zones above the piano.

**FIG. 16:**
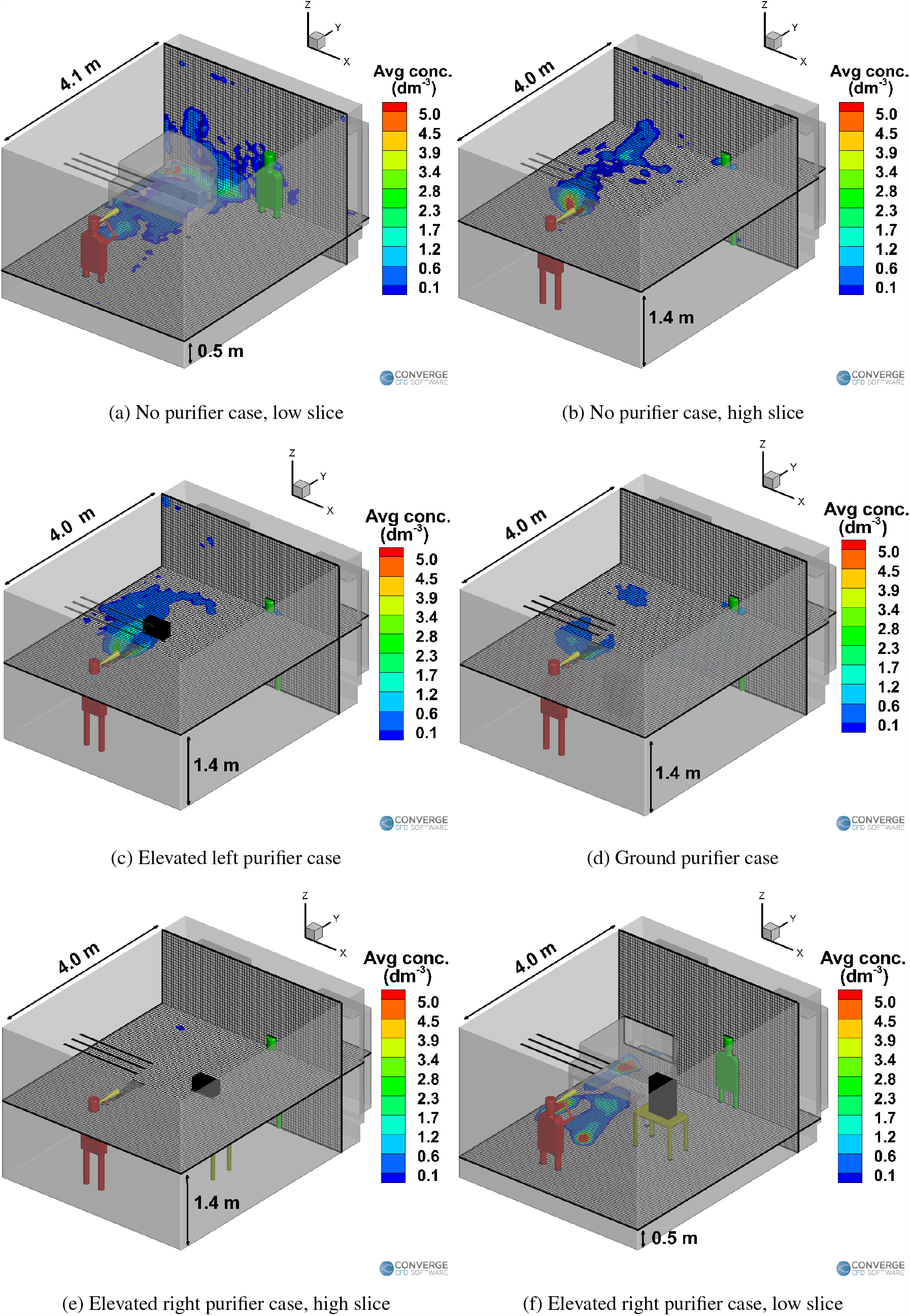
Time averaged airborne aerosol concentrations (per cubic decimeter) for the wind instrument (trombone) cases.

It is observed that the adding a purifier helps in reducing the airborne concentrations at the elevations of interest (a height of 1.4 m and a distance of 4.0 m from the front side). The purifier on the left side causes slightly more spreading of the aerosols onto the walls at the left (Fig. 16c) due to the airflow streamlines. The purifier on the ground further reduces the airborne aerosol concentration, as seen in Fig. 16d. This is because the purifier is kept in a location such that it offers the maximum removal of aerosols. The purifier on the right case is an interesting case; although it does reduce the aerosol concentrations at the elevations near the injector (i.e, a height of 1.4 m), there is actually a higher number of airborne aerosols when compared to any of the other case (including the no purifier case), as seen in Fig. 15a. Most of these airborne aerosols are in fact located in the region underneath the piano, as seen in Fig. 16f. This situation could be dangerous when the purifier is switched off, as these airborne aerosols might once again follow the natural recirculation streamlines underneath the piano and enter the space next to the instructor again. Moreover, this case offers zero removal of aerosols by the purifier, which again defeats the purpose of having a purifier in the first place.

In general, it is observed that the left side of the room experiences a higher concentration of airborne aerosols than the right side of the room. This result is consistent with the observations made earlier regarding the airflow field. Since the teacher is situated at a location slightly near the center-right side of the room, they are still exposed to a few airborne aerosols. We conclude that the teacher needs to be situated as close to the right side of the room as possible, since the risk of encountering airborne aerosols is significantly reduced there.

### B. Effect of injection rates: changing the instrument / using a mask / different modes of injection

In this section, the following cases are examined and compared: 1) student playing a wind instrument-a trumpet this time-inside a room (no purifier) with a teacher present for 11 minutes, 2) a student singing alone in a room wearing a surgical face mask for 11 minutes followed by a 25 minute break, and 3) a student playing a piano (wearing a cloth mask) for 11 minutes inside a room with a teacher. These are three typical scenarios encountered in a music classroom, and hence were chosen as the settings for this study. Although the cases are pertaining to a specific musical setting, the effects and analysis are not; they can be applied to any injection scenario in general. The different settings effectively just change the injection rate/flow streamlines, so that generalized observations on their influence can be made.

Figure 17 compares the temporal aerosol number profiles between the cases having different injection rates. First, the trombone case (Fig. 17a) and trumpet case (Fig. 17b) are compared with each other. Injection rates of 30 aerosols/s (trombone) and 100 aerosols/s (trumpet) were assumed based on the experimental measurements^52^, with both cases having airflow rates of 600 mL/s^68^. It is observed that the playing a trumpet around 13,581 aerosols (or 22.6% of the total 60,000 aerosols injected in the domain) to remain in the air, which is much higher compared to the trombone case (3,951 aerosols, or 21.9% of the 18,000 aerosols injected into the domain). Moreover, the number of deposited aerosols is also much higher for the trumpet case (46,416 aerosols, 77.4%) compared to the trombone case (14,049 aerosols, 78%). This consequently makes playing a trumpet riskier than playing a trombone. Interestingly, the number of airborne and deposited aerosols has roughly tripled in number for the trumpet case (compared to the trombone case), from 3,951 to 14,049, which is also the same scaling in the aerosol injection rate (which has roughly tripled from 30/s to 100/s).

**FIG. 17:**
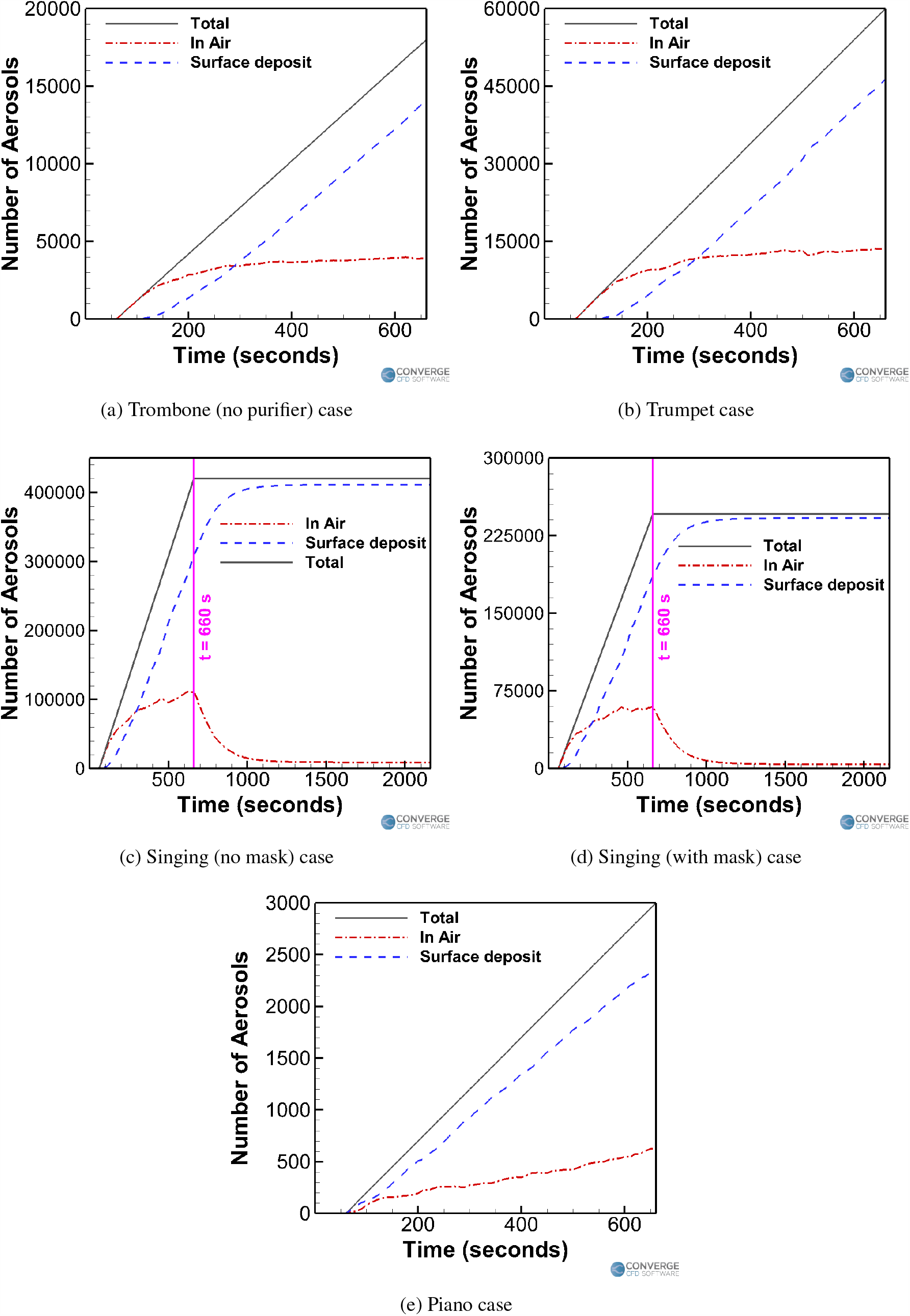
Effect of the injection rates on the temporal aerosol profiles.

Next, the student singing without a mask (Fig. 17c) and with a mask (Fig. 17d) are compared with each other. Injection rates of 700 aerosols/s (no mask) and 410 aerosols/s (with mask) were assumed^55^, with an airflow rate of 0.2 L/s^53,66^. For the first 11 minutes of simulation where the student continues to inject aerosols, it is observed singing with a mask drastically lowers the airborne aerosol number (see Fig. 17d) to 60,337 aerosols (or 24.5% of the total injected 246,000 aerosols), compared to the singing without a mask case (see Fig. 17c), which has a corresponding number of 111,943 aerosols (26.6% of the total of 420,000 aerosols). The number of surface deposited aerosols within the first 11 minutes have also decreased in the masked case (to 185,561 aerosols or 75.4%) compared to the non-masked case (307,957 aerosols or 73.3%). Similar to what was observed when comparing the trombone and trumpet cases, the airborne and deposited aerosol numbers (for the duration when the student continues to inject aerosols, i.e., 11 minutes) seem to have roughly halved when the injection rate was also roughly halved (from 700 to 410 aerosols/s). Once the student stops injecting aerosols, the remaining aerosols in the air are allowed to deposit or be removed. The final aerosol numbers for the masked singing case are 3,871 (1.7%) and 242,004 (98.3%) for the in air and surface deposited aerosols respectively, and 8,699 (2%) and 411,108 (97.8%) (in air and surface deposited aerosols, respectively) for the no mask singing case. Again, the final ratios of these numbers are roughly 1:1.7 for the singing with mask to singing without mask ratio, which again is similar to the 1:1.7 injection rate ratio of 410:700.

Lastly, the student playing a piano case is examined and compared (Fig. 17e) to the other cases. The student is just breathing normally, through a mask while playing the piano for a duration of 10 minutes. The aerosol injection rate is around 5 aerosols/s, assuming a 50% efficiency cloth mask^69^ with a normal aerosol breathing injection rate of 10 aerosols/s^52,55^. The exhaled airflow rate is around 0.1 L/s^53^. This case has the least amount of aerosols in the air (623 aerosols or 20.7% of the total 3,000 aerosols injected) and deposited aerosols (2,327 aerosols or 77.6%) compared to the other cases, making it the least dangerous scenario by a large margin.

Figure 18a shows the deposition of the aerosols onto the surfaces in the domain for the piano case. Major deposition occurs right on the student (Fig. 18b), due to the low air flow rate through the mask. Thus, the student’s clothes need to be thoroughly washed later, to remove the risk of spreading via contact. Further deposition occurs on the front of the piano, as well as the vent strip behind the piano. The deposition for the piano case is very less, compared to any of the singing (Fig. 18d) or wind instrument cases (trombone, Fig. 18c).

**FIG. 18:**
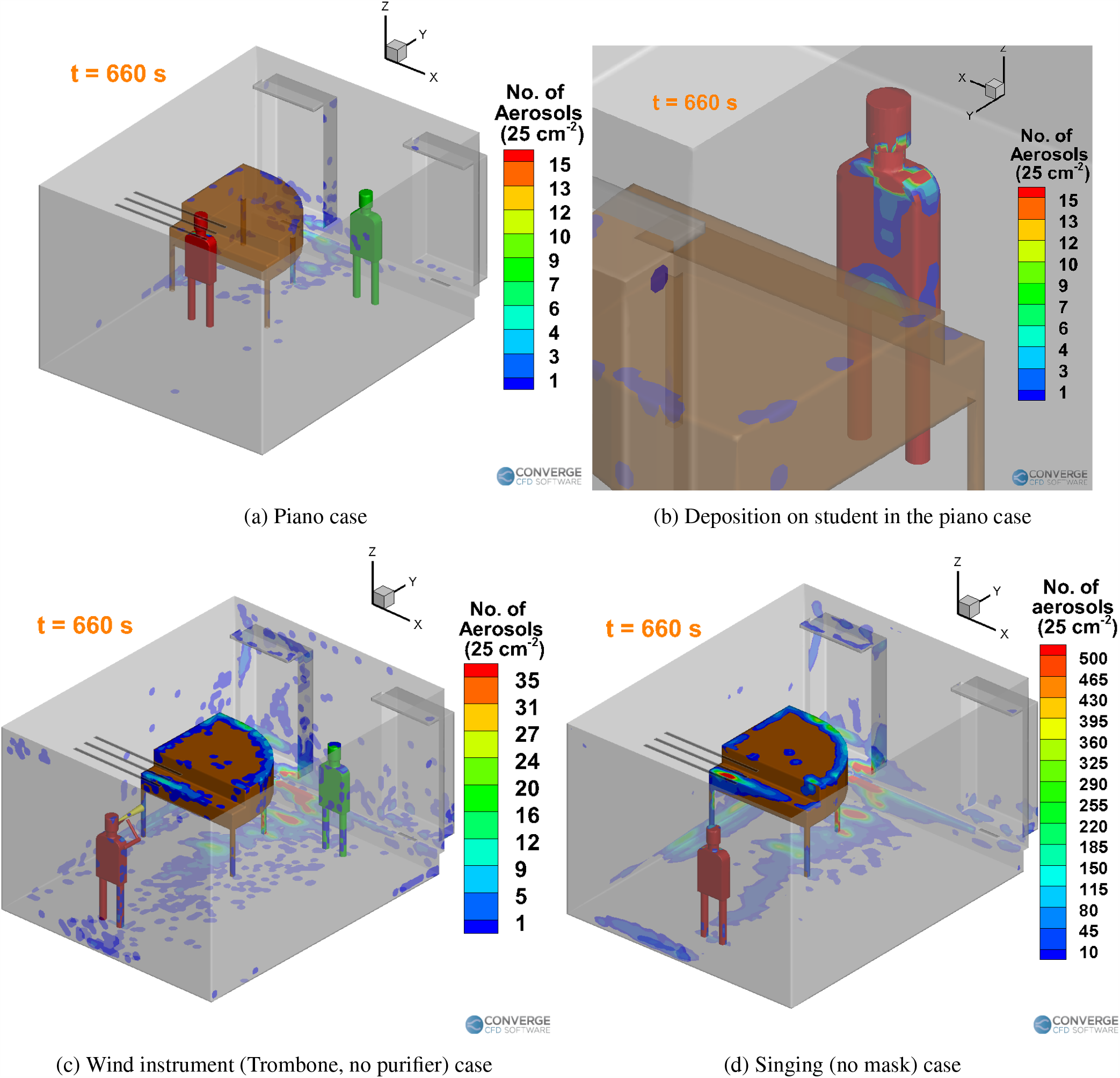
Comparison of the deposition of aerosols inside the domain between the three types of musical sessions.

### C. Summarizing the overall trends

Figure 19 shows the trends of the airborne and deposited aerosol numbers when comparing the two types of effects - the effect of a purifier (Fig. 19a) and varying the injection rate (Fig. 19b). Although the numbers may change depending on the simulation time, the trends will hold good for any simulation time larger than the initial transient periods specified in previous sections.

**FIG. 19:**
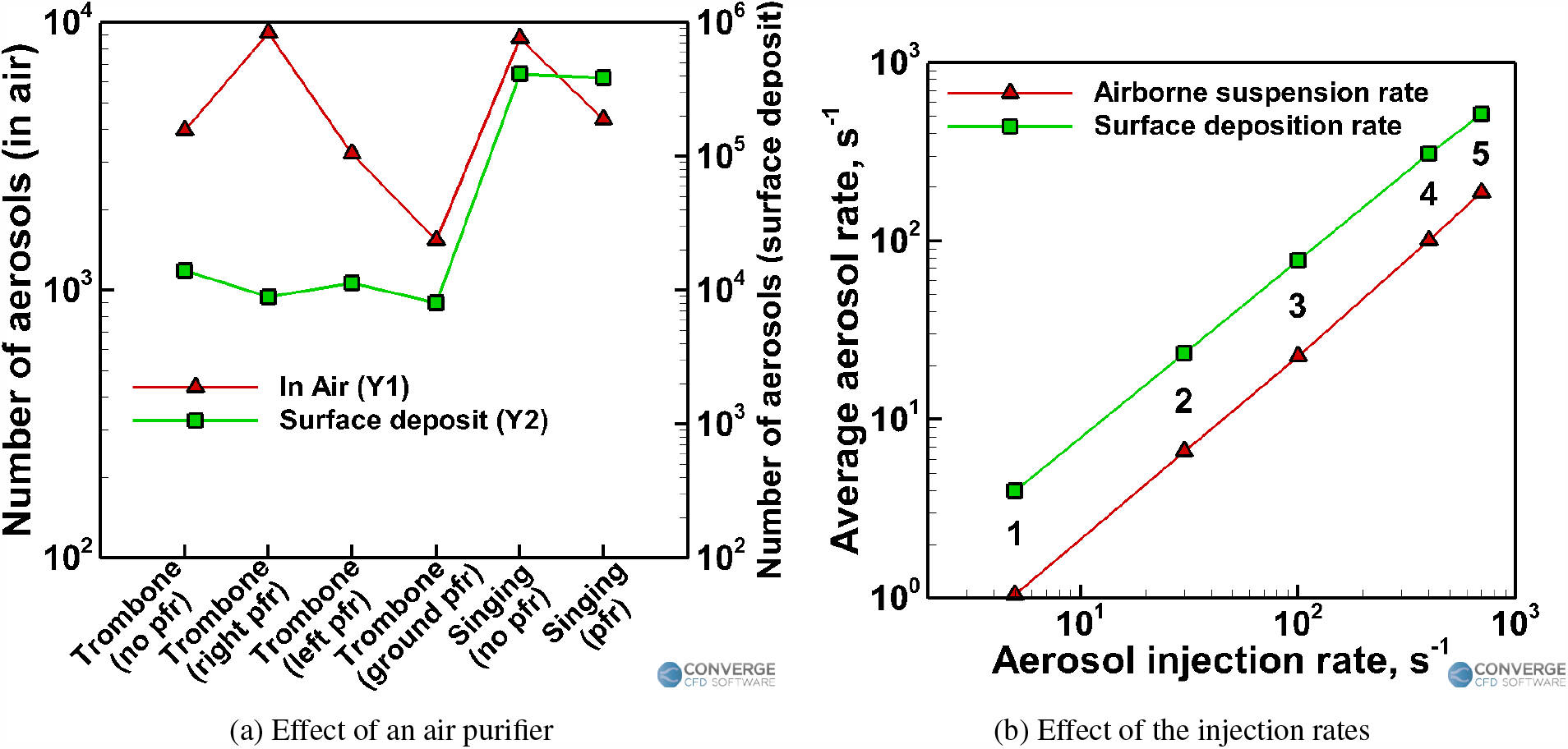
Summarizing the observed trends.

Figure 19a shows the total number of aerosols remaining in the domain by the end of the simulation (in the instruments case, the simulation ends after the injection stops. In the singing cases, the simulation ends after 25 minutes of idle time following the 10 minutes of injection). The advantage of using purifiers is heavily dependent on the location of the purifier. As explained earlier, improper placement of the purifier might even make the situation worse, as is evident from the purifier on the right side case (where the resulting airborne aerosol number is almost thrice that of the case without a purifier, and almost ten times that of the case with a purifier on the ground, Fig. 19a). Using a purifier almost halves the airborne aerosol number in the singing case. Smart placement of purifiers can thus yield a huge benefit in terms of reducing the airborne aerosol number. The optimal location for purifier placement highly depends on the geometry of the domain and the flow parameters. The first and most important step in any general domain is to identify the region where placing a purifier would yield a positive benefit, and not a negative effect. This region of benefit is usually the area right in front of the injector (i.e., the infected person, assumed to be the student here), and the vicinity around the injector. It would be advisable to place the purifier in the front of the injector in any domain. For this case, The elevation of the purifier is advised to be kept closer to the ground (could be a few centimetres off the ground as well) rather than at an high elevation, since the aerosols don’t ballistically travel forward at the same elevation of the injector (like large droplets). These aerosols follow the air streamlines and depending on those streamlines, an optimal height can be ascertained. Placing the purifier farther away from the injector reduces the effectiveness of the purifier, and in some cases worsens the situation. Thus, placing a purifier near the person whom you want to protect (in this case, the teacher) is not advisable. The streamlines from the purifier might pull aerosols which are far away from the teacher towards the purifier (and hence towards the teacher), which could be dangerous. Interestingly, the number of deposited aerosols is more or less unaffected by the presence of purifiers, as seen from both the wind instrument and the singing case (Fig. 19a). This is because a large portion of the aerosols are still deposition dominated; the aerosols get deposited at the various surfaces (such as the piano, the underside of the piano, the vent strip, the left side window and the left side wall) since the solid objects/surfaces are located in spots where they directly obstruct the aerosol flow paths. Hence, these surfaces are still going to experience similar levels of deposition even if the flow patterns slightly change. The recirculation zones starting from underneath the piano and moving upwards towards the ceiling on the back side of the piano also remain consistent among all the cases, which makes the deposition patterns in these regions consistent. The deposition rate also seems to be higher than the removal rate of the aerosols. Hence, while the deposition more or less occurs at a similar rate (on the afore-mentioned regions) among similar cases, the removal vastly differs because of the purifier’s effect on the streamlines.

According to Fig. 19b, the injection rate is observed to vary linearly with both the average rate of aerosols being suspended in the air as well as the average rate of aerosols being deposited in the domain. The average rate here is defined as the total number of aerosols present in air or deposited onto surfaces (which is shown in Table IV) divided by the total injection time (10 minutes in this study).

**TABLE IV:** The total number of aerosols remaining in the domain (after 10 minutes of injection).

| CASE | Type of injection mode | Aerosol injection rate ($s^{-1}$) | Number of aerosols (in air) | Number of aerosols (deposited) |
| --- | --- | --- | --- | --- |
| 1 | Playing a piano | 5 | 623 | 2,377 |
| 2 | Playing a trombone | 30 | 3,951 | 14,049 |
| 3 | Playing a trumpet | 100 | 13,581 | 46,416 |
| 4 | Singing with a mask | 410 | 60,337 | 185,561 |
| 5 | Singing without a mask | 700 | 111,943 | 307,957 |

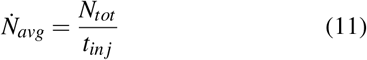

where 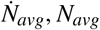 and *t*_*inj*_ refer to the average aerosol rate, total number of aerosols and time of injection respectively. The linear correlation between the injection rate and the average aerosol rate from this study is shown below:

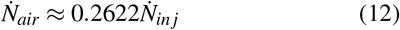

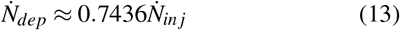

where 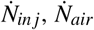 and 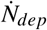 refer to the number of aerosols injected per second, average airborne aerosols suspension rate and the average aerosol deposition rate, respectively. The above observations suggest that a linear trend can be expected between the injection rate and the number of airborne/deposited aerosols in a domain within a given time frame.

## IV. CONCLUSION

In this study, the effects of portable purifiers and aerosol injection rates were analyzed for different settings typically observed in a music classroom. The three categories of cases (chosen based on the typical classroom scenarios in a music school) were (a) a student singing alone (with and without a purifier/mask), (b) a student playing a wind instrument (trombone or trumpet) in the presence of a teacher (with and without a purifier), and (c) a student playing the piano (wearing a mask) in the presence of a teacher. Although these cases were chosen in interest of the University of Minnesota School of Music, the resulting analysis and observations made here could be useful for general aerosol injection scenarios. The cases here effectively vary the aerosol injection rates and/or airflow streamlines, which are not specific to a musical setting.

Using purifiers help in achieving ventilation rates as suggested by WHO and CDC guidelines, since the in-built HVAC ventilation rates of the building may not be sufficient to achieve the desired aerosol removal rates. It was observed that using a purifier aids in improving the natural ventilation (through the building vents), which further helps in achieving removal times within the CDC prescribed values. This fact helps in arriving at an effective break period of 25 minutes between class sessions, where the airborne aerosol removal using a purifier is almost 97%. Since the number of airborne aerosols fluctuate about a mean value after an initial transient period (around 5 to 9 minutes), the break period will apply to any typical small classroom duration.

The effect of purifiers was found to offer significant benefits, provided the purifiers were placed in proper locations, offering orders of magnitude higher rates of aerosol removal compared to cases having no purifiers (where the removal might even be close to zero). Placing the purifiers close to the injector, and specifically in the path of aerosol injection/transport could offer positive benefits. The purifier should also preferably be kept in a position where the exhaust flow aids in the natural recirculation of the room. This can be found in the purifier on the ground case where the exhaust airflow from the purifier travels underneath the piano and follows the natural recirculation zone above. Placing the purifier in a location where the exhaust flow significantly changes the airflow streamlines in the domain (e.g., purifier on the left or right side at an elevation), and causes mixing in the domain may cause more spreading of the aerosols (although some reduction in total airborne aerosols may still be achieved). The study therefore advises that the purifier be kept close to the injector, and away from the individuals whom you want to protect. This could apply even to a case with multiple purifiers.

Finally, an almost linear correlation between the injection rate and the number of aerosols remaining in the air and deposited onto surfaces was observed. This detail could predict similar quantities when a different injection rate is used, without the need for simulating the entire domain again.

## Data Availability

The data that support the findings of this study are available from the corresponding author upon reasonable request.

## ACKNOWLEDGMENTS

This work was supported by the University of Minnesota School of Music. The authors would like to thank Peter Remiger and Michael Kim from the School of Music for their valuable inputs and helpful discussions. Convergent Science provided CONVERGE licenses and technical support for this work.

